# EEG functional connectivity as a prognostic biomarker of adaptive function in autistic people

**DOI:** 10.1101/2025.05.17.25327836

**Authors:** Chirag Mehra, Petroula Laiou, Pilar Garcés, Joshua B Ewen, Eva Loth, Mark H Johnson, Luke Mason, Emily JH Jones, Tony Charman, Thomas Bourgeron, Jan Buitelaar, Michael Absoud, Mark P Richardson, Declan Murphy, Jonathan O’Muircheartaigh

**Affiliations:** Department of Forensic and Neurodevelopmental Sciences, Institute of Psychiatry, Psychology and Neuroscience, King’s College London, London, UK; Wolfson Neurodisability Service, Great Ormond Street Hospital for Children NHS Foundation Trust, London, UK; Department of Basic and Clinical Neuroscience, Institute of Psychiatry, Psychology and Neuroscience, King’s College London, London, UK; Department of Biostatistics & Health Informatics, Institute of Psychiatry, Psychology and Neuroscience, King’s College London, London, UK; Roche Pharma Research and Early Development, Neuroscience and Rare Diseases, Roche Innovation Center Basel, F. Hoffmann–La Roche Ltd., Basel, Switzerland; Developmental & Behavioral Pediatrics, Ann & Robert H. Lurie Children’s Hospital of Chicago, Chicago, Illinois, USA; Department of Pediatrics at Northwestern University Feinberg School of Medicine, Chicago, Illinois, USA; Department of Psychology, University of Cambridge, Cambridge, UK; Department of Child and Adolescent Psychiatry, Institute of Psychiatry, Psychology and Neuroscience, King’s College London, London, UK; MRC Centre for Developmental Neurobiology, Institute of Psychiatry, Psychology and Neuroscience, King’s College London, London, UK; Department of Psychology, Institute of Psychiatry, Psychology and Neuroscience, King’s College London, London, UK; Human Genetics and Cognitive Functions, Institut Pasteur, UMR3571 CNRS, Université de Paris, 75015 Paris, France.; Department of Medical Neuroscience, Donders Institute for Brain, Cognition and Behaviour, Radboud University Medical Centre, Nijmegen, Netherlands.; Department of Women and Children’s Health, Faculty of Life Sciences and Medicine, School of Life Course Sciences, King’s College London, London, UK; Children’s Neurosciences, Evelina London Children’s Hospital at Guy’s and St Thomas’ NHS Foundation Trust, London, UK; Department of Early Life Imaging, School of Biomedical Engineering and Imaging Sciences, King’s College London, UK

**Author notes:** Correspondence to: Chirag Mehra, Department of Forensic and Neurodevelopment Sciences, Institute of Psychiatry, Psychology and Neuroscience, King’s College London, 16 De Crespigny Park, London, SE5 8AF, UK.

**Keywords:** autism, functional connectivity, adaptive function, prognostic biomarker, EEG

## Abstract

Many autistic people have challenges with adaptive function, impacting education, employment and independent-living goals. Adaptive function outcomes of autistic people vary considerably, which makes planning for future needs challenging. Here, using a developmentally sensitive approach, we investigated if cortico-cortical functional connectivity – a core neurobiological feature that differs in autism – could predict longitudinal changes in adaptive function in autistic people. Using electroencephalography in 150 autistic and 159 non-autistic participants aged 6-31 years, we investigated if mean degree and network organisation (small-world index) predict longitudinal changes in adaptive function over 19-months. We found that small-world index significantly predicted changes in adaptive function in autistic people across the entire age-range. Predictive performance was best for autistic youth (15-24-year-olds), where mean degree and small-world index explained 21% and 30% of additional variance in outcomes, respectively, outperforming measures of intelligence and autistic features. In categorising binary (improved versus not-improved) outcomes, the model containing mean degree had an AUC of 0.84 [95% CI: 0.71-0.97] in 15-24-year-olds, while that containing small-world index had an AUC of 0.76 [95% CI: 0.63-0.89] across the 6-31-year age-range. Both metrics demonstrated properties desired in prognostic biomarkers: high test-retest reliability and convergence with underlying biology (significant associations with genetic variation in brain volume-related genes). Thus, we demonstrate the first evidence that electroencephalography-derived functional connectivity metrics show promise as prognostic biomarkers of adaptive function in autistic people. Potential precision-medicine applications include stratifying participants in clinical trials and identifying those at risk of declining function in clinical settings.

## Introduction

A minority of autistic adults work full-time (12-27%), are enrolled in or have completed postsecondary education (39%) or live independently (5-12%; 1–3). Consequently, identifying approaches to support function in these domains is a research priority of autistic people (4,5). The neurocognitive construct most strongly associated with these abilities in autistic people is adaptive function (1,3,6–8): the application of one’s abilities to navigate the everyday environment, including in a self-sufficient manner (9). Indeed, adaptive function has been a key outcome measure in autism clinical trials (10,11). However, there is substantial variation in how adaptive function abilities of autistic people change over time (12,13). This poses a challenge in helping autistic young-people and adults plan requirements for future support or independence, and risks concealing important subgroup effects in clinical trials (14), highlighting a need for precision-medicine approaches.

Current behavioural predictors of improvements in adaptive function in autistic people – higher intelligence (1,15,16), higher baseline adaptive function (1,16) and speech abilities (16) – leave most of the variance in adaptive function outcomes unexplained (9,15,17–19). Existing putative neurobiological predictors – including the speed of neural response to faces (the N170 event-related potential; 14) and cortical structural differences (20) – are either limited in the domains of adaptive function outcomes they predict (14) and/or only explain a small proportion of additional variance (6% each) in outcomes (14,20). This highlights the need for better prognostic biomarkers.

Several studies have demonstrated that brain connectivity is different in autistic compared to non-autistic people (21–27). Both the amount (often quantified as mean degree; 21,28,29) and organisation (30–33) of functional connectivity in large-scale brain networks have been associated with the extent of autistic social-communication and behavioural traits. An important feature of brain-networks across species is their small-world organisation (34–36), which facilitates specialised (segregated) and co-ordinated (integrated) function within and across neural systems (34,37). Altered small-world organisation has been found in autism (30–32). Therefore, these functional connectivity metrics may show promise as prognostic biomarkers for adaptive function in autistic people.

However, previous functional connectivity findings in autistic people have been heterogenous (22,24,38–40), limiting progress in biomarker discovery. Considering age-effects in autism research is especially important given that autism is associated with altered rates of white matter (22,33,41–43), brain volume (44) and behavioural (45) maturation. Yet, most prior studies have inadequately accounted for developmental stage (40), possibly contributing to heterogenous findings.

Here, we investigated the performance of functional connectivity-derived mean degree (MD) and small-world index (SWI) as candidate prognostic biomarkers for adaptive function outcomes in autistic people. We explicitly studied age effects in a large dataset comprising participants aged 6-31 years, hypothesising that the extent to which MD/SWI predicts adaptive function outcomes differs by developmental stage. A candidate prognostic biomarker needs to be associated with underlying biological processes, statistically associated with an outcome, quantifiable in real-world settings and highly reliable (46–48). We assessed convergence of functional connectivity metrics with biological processes by quantifying associations with polygenic variation, as autism (49,50) and functional connectivity (51–53) are both highly heritable. We explored polygenic variation in autism- and brain-volume-related genes, the latter motivated by findings of altered brain volume (20,44) and arguments that brain volume differences underly functional connectivity differences in autistic people (54). We measured neural activity using electroencephalography (EEG) as it is widely available in clinical settings and cost-effective (55). EEG-based functional connectivity also reflects the mechanisms underlying neural interactions (56–58). Finally, we quantified the reliability of MD and SWI.

## Participants and Methods

Details are included in the Supplementary Methods section.

### Ethics approval and consent to participate

This study was approved by ethics review boards at each study site, as detailed in Loth *et al*., (59). The ethics committees were: 1) London-Central and Queen Square Health Research Authority Research Ethics Committee in the United Kingdom (13/LO/1156); 2) Regio Arnhem-Nijmegen (Radboud University Medical Centre Institute Ensuring Quality and Safety Committee on Research Involving Human Subjects Arnhem-Nijmegen) and 3) Radboud universitair medisch centrum Instituut Waarborging Kwaliteit en Veiligheid Commissie Mensgebonden Onderzoek in the Netherlands (2013/455); 4) UMM Universitätsmedizin Mannheim, Medizinische Ethik Kommission II (UMM University Medical Mannheim, Medical Ethics Commission II) in Germany (2014-540N-MA); and 5) Universita Campus Bio-Medica De Roma Comitato Etico (University Campus Bio-Medical Ethics Committee De Roma) in Italy (18/14 PAR ComET CBM).

Participant informed consent was obtained in keeping with the Declaration of Helsinki. All methods were performed in accordance with the relevant guidelines and regulations, where applicable.

### Participants

We included 150 autistic (27% female) and 159 non-autistic (33% female) participants, aged 6-31 years, from five sites of the AIMS-2-TRIALS Longitudinal European Autism Project (59,60), Table 1a. Autistic participants had an existing clinician-made diagnosis. Included participants had at least one 20-second epoch of good-quality eyes-closed resting-state EEG data and a good-quality T1 structural MRI image (for source reconstruction). Given that mental health and other developmental conditions commonly co-occur with autism (64), we did not exclude participants with these conditions. We excluded participants with neurological conditions, gross structural brain lesions, diagnosed intellectual disability, a full-scale IQ < 75 or significant systemic disease.

**Table 1.**
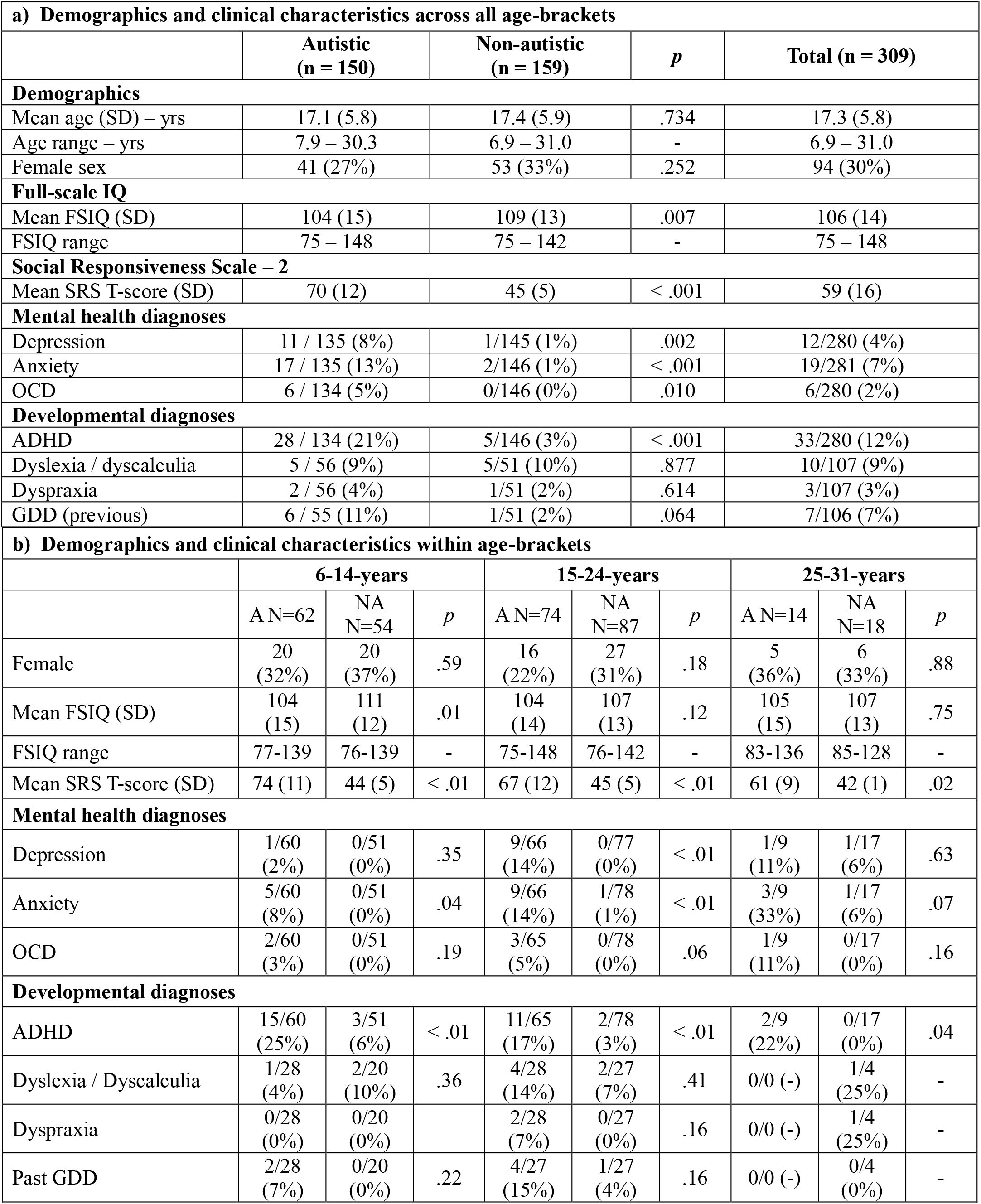
Demographics and clinical characteristics of participants. FSIQ = full-scale IQ. OCD = obsessive compulsive disorder. ADHD = attention deficit / hyperactivity disorder. GDD = global developmental delay. A = autistic, NA = non-autistic.

| a) Demographics and clinical characteristics across all age-brackets |  |  |  |  |  |  |  |  |  |
| --- | --- | --- | --- | --- | --- | --- | --- | --- | --- |
|  | Autistic<br>(n = 150) |  |  | Non-autistic<br>(n = 159) |  |  | *p* | Total (n = 309) |  |
| Demographics |  |  |  |  |  |  |  |  |  |
| Mean age (SD) – yrs | 17.1 (5.8) |  |  | 17.4 (5.9) |  |  | .734 | 17.3 (5.8) |  |
| Age range – yrs | 7.9 – 30.3 |  |  | 6.9 – 31.0 |  |  | - | 6.9 – 31.0 |  |
| Female sex | 41 (27%) |  |  | 53 (33%) |  |  | .252 | 94 (30%) |  |
| Full-scale IQ |  |  |  |  |  |  |  |  |  |
| Mean FSIQ (SD) | 104 (15) |  |  | 109 (13) |  |  | .007 | 106 (14) |  |
| FSIQ range | 75 – 148 |  |  | 75 – 142 |  |  | - | 75 – 148 |  |
| Social Responsiveness Scale – 2 |  |  |  |  |  |  |  |  |  |
| Mean SRS T-score (SD) | 70 (12) |  |  | 45 (5) |  |  | < .001 | 59 (16) |  |
| Mental health diagnoses |  |  |  |  |  |  |  |  |  |
| Depression | 11 / 135 (8%) |  |  | 1/145 (1%) |  |  | .002 | 12/280 (4%) |  |
| Anxiety | 17 / 135 (13%) |  |  | 2/146 (1%) |  |  | < .001 | 19/281 (7%) |  |
| OCD | 6 / 134 (5%) |  |  | 0/146 (0%) |  |  | .010 | 6/280 (2%) |  |
| Developmental diagnoses |  |  |  |  |  |  |  |  |  |
| ADHD | 28 / 134 (21%) |  |  | 5/146 (3%) |  |  | < .001 | 33/280 (12%) |  |
| Dyslexia / dyscalculia | 5 / 56 (9%) |  |  | 5/51 (10%) |  |  | .877 | 10/107 (9%) |  |
| Dyspraxia | 2 / 56 (4%) |  |  | 1/51 (2%) |  |  | .614 | 3/107 (3%) |  |
| GDD (previous) | 6 / 55 (11%) |  |  | 1/51 (2%) |  |  | .064 | 7/106 (7%) |  |
| b) Demographics and clinical characteristics within age-brackets |  |  |  |  |  |  |  |  |  |
|  | 6-14-years |  |  | 15-24-years |  |  | 25-31-years |  |  |
|  | A N=62 | NA<br>N=54 | *p* | A N=74 | NA<br>N=87 | *p* | A N=14 | NA<br>N=18 | *p* |
| Female | 20<br>(32%) | 20<br>(37%) | .59 | 16<br>(22%) | 27<br>(31%) | .18 | 5<br>(36%) | 6<br>(33%) | .88 |
| Mean FSIQ (SD) | 104<br>(15) | 111<br>(12) | .01 | 104<br>(14) | 107<br>(13) | .12 | 105<br>(15) | 107<br>(13) | .75 |
| FSIQ range | 77-139 | 76-139 | - | 75-148 | 76-142 | - | 83-136 | 85-128 | - |
| Mean SRS T-score (SD) | 74 (11) | 44 (5) | < .01 | 67 (12) | 45 (5) | < .01 | 61 (9) | 42 (1) | .02 |
| Mental health diagnoses |  |  |  |  |  |  |  |  |  |
| Depression | 1/60<br>(2%) | 0/51<br>(0%) | .35 | 9/66<br>(14%) | 0/77<br>(0%) | < .01 | 1/9<br>(11%) | 1/17<br>(6%) | .63 |
| Anxiety | 5/60<br>(8%) | 0/51<br>(0%) | .04 | 9/66<br>(14%) | 1/78<br>(1%) | < .01 | 3/9<br>(33%) | 1/17<br>(6%) | .07 |
| OCD | 2/60<br>(3%) | 0/51<br>(0%) | .19 | 3/65<br>(5%) | 0/78<br>(0%) | .06 | 1/9<br>(11%) | 0/17<br>(0%) | .16 |
| Developmental diagnoses |  |  |  |  |  |  |  |  |  |
| ADHD | 15/60<br>(25%) | 3/51<br>(6%) | < .01 | 11/65<br>(17%) | 2/78<br>(3%) | < .01 | 2/9<br>(22%) | 0/17<br>(0%) | .04 |
| Dyslexia / Dyscalculia | 1/28<br>(4%) | 2/20<br>(10%) | .36 | 4/28<br>(14%) | 2/27<br>(7%) | .41 | 0/0 (-) | 1/4<br>(25%) | - |
| Dyspraxia | 0/28<br>(0%) | 0/20<br>(0%) |  | 2/28<br>(7%) | 0/27<br>(0%) | .16 | 0/0 (-) | 1/4<br>(25%) | - |
| Past GDD | 2/28<br>(7%) | 0/20<br>(0%) | .22 | 4/27<br>(15%) | 1/27<br>(4%) | .16 | 0/0 (-) | 0/4<br>(0%) | - |

#### Age-groups

For the age-group analyses, participants were split into 3 age-groups (Table 1b): children (6-14 years, n = 116), youth (15-24 years, n = 161), and adults (25-31 years, n = 32). Youth was defined as per the United Nations World Health Organisation (65,66). This age-based definition aligns with a period of disruptive maturation of functional connectivity, in contrast to the conservative maturation seen in younger and older ages (67,68).

#### Behavioural measures

We measured adaptive function in autistic people at two timepoints (T1 and T2) using the Vineland Adaptive Behaviour Scale (VABS), second edition (69). The VABS is a semi-structured interview, which we conducted with participants’ parent or carer. Adaptive function across the domains of communication, daily living and socialisation skills was quantified by the adaptive behaviour composite score (ABC), standardised for age (mean 100, SD 15). Higher scores correspond with more age-normed adaptive behaviours. We calculated longitudinal changes in adaptive function (ΔABC) by subtracting T2 from T1 ABC scores, as done previously (14,20).

The age-standardised ‘T-scores’ of the Social Responsiveness Scale-Second Edition (SRS-2; 70) were used as trait measures of autistic behaviours. Higher T-scores correspond with more autistic behaviours. We used the parent-reported instead of self-reported T-scores in our main analysis, as we had a larger sample size with the former (n = 118 versus n = 76) and the two measures are not interchangeable (71), Fig. S1. Intellectual ability was assessed using the Wechsler Abbreviated Scales of Intelligence-Second Edition (72), or shortened versions of the German, Dutch or Italian Wechsler Intelligence Scale for Children, third or fourth editions (73,74) or Wechsler Adult Intelligence Scale third or fourth editions (75,76).

### Electroencephalography methods

Four segments of 30-second, eyes-closed recordings were captured using 60–70 EEG electrodes (site dependent). We transformed EEG timeseries from sensor space to source space using individual-participant T1-weighted MRI images and a linearly constrained minimum variance beamformer (77), as per Garcés *et al.* (78).

We chose 20 second eyes-closed resting-state EEG epochs to produce functional connectivity matrices. This epoch length was a compromise between maximising the number of included participants and maximising network stability; EEG-functional-connectivity-derived brain-network organisation has been shown to be stable at epoch lengths of 10-12 seconds and above (79,80). We filtered signals between 6-9 Hz (‘low-alpha’).

#### Functional connectivity network analyses

For each 20-second epoch, we calculated functional connectivity between brain-regions using the phase locking value (PLV; 81). Scalp-EEG-derived PLV has high test-retest reliability, concordance with its underlying structural connectivity and is sensitive to age-related connectivity changes (82).

We created weighted, undirected functional connectivity matrices with 68 nodes. Each node corresponded to a cortical region of the Desikan-Killiany atlas (83), while the edges were the PLV between pairs of nodes. The network metrics of interest were the mean degree and the small-world index, calculated using the brain connectivity toolbox (84).

The degree of each node was averaged over 68 nodes to obtain the mean degree, *MD*:

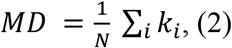

where *k_i_* denotes the number of edges of node *i* and *i* = 1, …, *N* = 68.

We used a modified (for weighted networks) Humphries and Gurney (85) definition of the weighted SWI (86). The weighted SWI, *σ*, was calculated by dividing the normalised weighted clustering coefficient, *C*′, by the normalised weighted pathlength, *L*′:

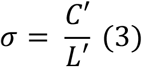

If a participant had more than one usable 20-second EEG epoch, MD and weighted SWI were calculated for each epoch. Each participant had 1-4 usable epochs. The per-participant connectivity metrics used in analyses were calculated from the mean of 1-4 epochs, as the mean of multiple measures is more reliable than a single measure (87).

#### Test-retest reliability

We quantified the intrasession test-retest reliability between two epochs recorded between 1-3 minutes of each other. Reliability was calculated for metrics derived from one 20-second epoch using interclass correlation coefficient (ICC (2,1)) and metrics derived from the mean of two 20-second epochs using ICC(2,k). Results were interpreted as per Ku and Li (88).

#### Genetics

We calculated polygenic likelihood scores (PGS) using blood samples. Using reference genome-wide association study summary statistics, PGS were produced for autism (89) and brain volume (90). A more positive brain volume PGS is associated with more alleles for large brain volume.

#### General linear models predicting longitudinal changes in adaptive function

Our primary aim was to investigate if functional connectivity metrics can predict longitudinal changes in ΔABC and if prediction ability varies by age. Longitudinal ABC data was available for 40 autistic children (6-14-years), 38 autistic youth (15-24-years) and 3 autistic adults (25-31-years), Fig S2. Therefore, we omitted the adults from the age-bracket analyses predicting ΔABC.

We used general linear models to predict ΔABC, adjusting them for T1 ABC, participant age and an age x T2 – T1 (follow-up interval) interaction term (as per Pretzsch *et al.* (20) – as the rate of behavioural change differs by developmental stage; 45). We also used these models to determine the presence of significant age x group interaction effects and adjust for the effect of nuisance covariates (e.g. site and sex). We retained interaction effects when *p* < .05 for the interaction term (91). We selected general linear models by comparing the Akaike information criterion (AIC) and the adjusted R^2^ between models with different nuisance co-variates. We evaluated the out-of-sample generalisability of the models using the connectome-based predictive modelling protocol (92).

In models where there was a main effect of functional connectivity metric on ΔABC, we explored the ability of our models to predict categorical ΔABC on a per-participant basis, using leave-one-out cross-validation. We used an ΔABC cutoff of 6 to define improving, worsening or static outcomes. This cutoff was based on evidence that ΔABC greater than ±2.7-8 (93,94) in autistic people without intellectual disability are minimal clinically-important differences and are unlikely (*p* < .05) to occur by chance given the standard error of measurement of the ABC (93,95,96). We assessed if predictive ability was better than chance using permutation analyses. Then, to illustrate predictive performance using the area under the receiver operated characteristic curve (AUC), sensitivity and specificity, we binarized outcomes to increased (≥ 6) or not-increased (< 6) ABC scores. 95% confidence intervals for AUC were quantified using bootstrapping.

Finally, we illustrated the relationship between effect size and age *without* pre-defined age brackets. We plotted a moving estimated coefficient of the functional connectivity metric predicting ΔABC as a function of a moving average participant age, as per O’Muircheartaigh *et al.* (97). To obtain estimated coefficients, we performed identical general linear models in participant groups of n = 30, ordered by participant age, increasing by 5 participants at a time.

## Code availability

The code used for these analyses can be found here https://github.com/cmehra- brain/EEG_FC_autism and is under a BSD-3-Clause license.

## Results

### EEG epochs

There were no differences between the number of analysable EEG epochs per participant between autistic and non-autistic people (*W* = 11362, *p* = .66). Participants with more usable epochs had a greater mean age *H*(3) = 8.9, *p* = .03 (Fig. S3).

### Cross-sectional group differences in functional connectivity

We quantified the extent of cortico-cortical functional connectivity using mean degree. Age-associated increases in MD were slower in autistic than in non-autistic people (β = -0.19, SE = 0.10, *p* = .045, η^2^ = 0.01), Fig. 1a, Table S1. Compared to non-autistic people, MD was no different in autistic 6-14-year-olds (β = 0.11, SE = 0.18, *p*_FDR_ = .52) and 25-31-year-olds (β = -0.32, SE = 0.45, *p*_FDR_ = .52) but lower in autistic 15-24-year-olds (β = -0.46, SE = 0.15, *p*_FDR_ = .005), Fig. 1c. In autistic people aged 6-31-years, the presence of more parent-reported autistic features (SRS T-score) correlated with lower MD, even when adjusting for site effects (partial *r* = -.31, n = 118, *p* < .001), Fig. 1e.

**Figure 1.**
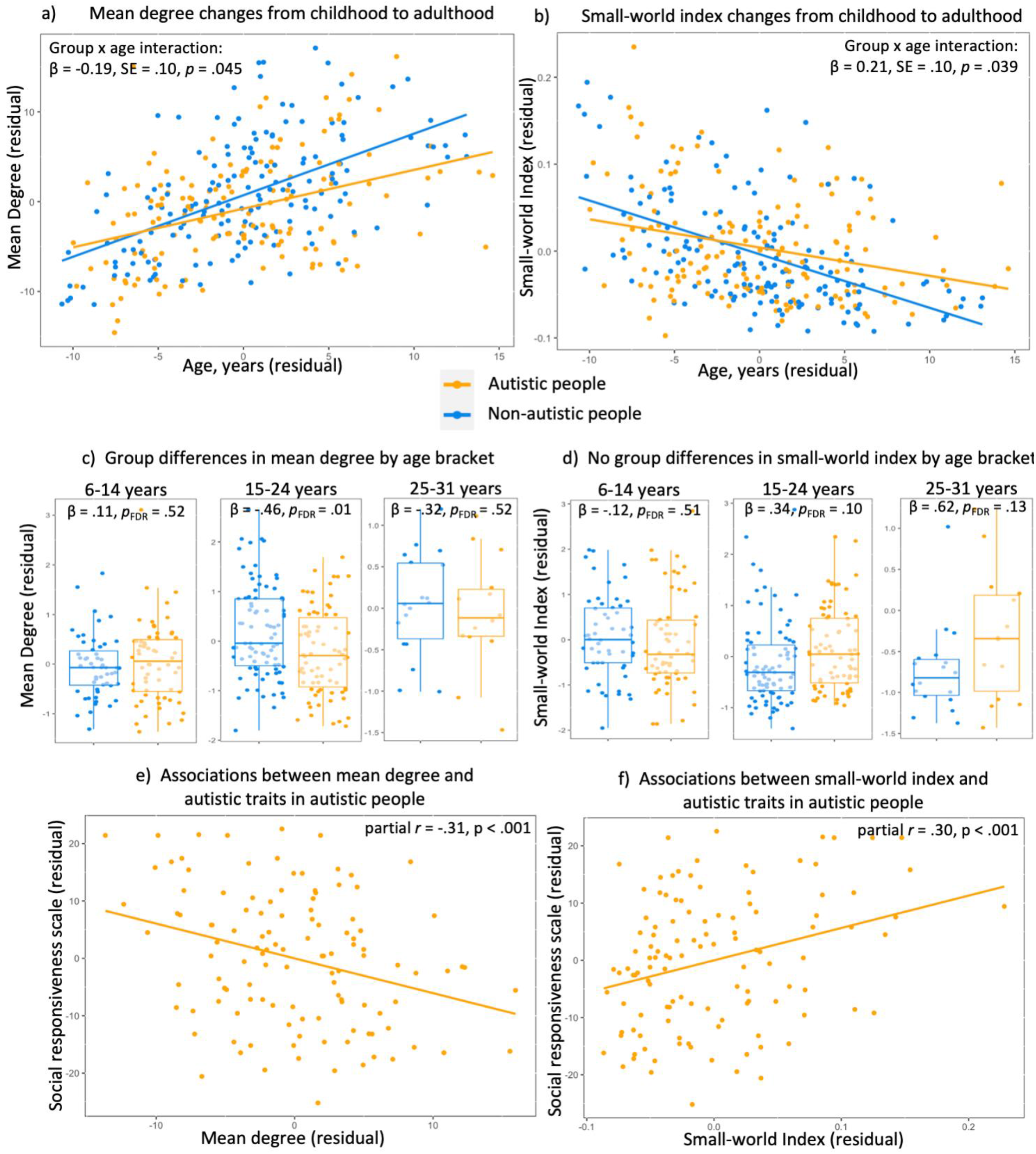
Cross-sectional associations between functional connectivity metrics and autism. In both autistic and non-autistic people, **a)** mean degree significantly increased and **b)** small-world index significantly decreased with age from childhood to adulthood. **a-b)** There were significant group x age interactions in mean degree and small-world index: both changed more slowly from childhood to adulthood in autistic people (n = 309)**. c)** Mean degree was no different in autistic 6-14-year-olds (n = 116) or autistic 25-31-year-olds (n = 32) but was lower in autistic 15-24-year-olds (n = 161), compared to non-autistic people. **d)** Small-world index did not differ by group in any age-group. In autistic people, **e)** lower mean degree and **f)** higher small-world index was associated with more autistic features (n = 118).

Age-associated decreases in SWI were slower in autistic than in non-autistic people (β = 0.21, SE = 0.10, *p* = .039, η^2^ = 0.01), Table S2. We found no differences in SWI in autistic 6-14-year-olds (β = -0.12, SE = 0.18, *p*_FDR_ = .51), 15-24-year-olds (β = 0.34, SE = 0.16, *p*_FDR_ = .10) or 25-31-year-olds (β = 0.62, SE = 0.34, *p*_FDR_ = .13) compared to non-autistic people, Fig. 1d.

Within autistic people aged 6-31-years, there was a positive correlation between parent-reported SRS T-score and SWI, even when adjusting for site effects (partial *r* = .30, n = 118, *p* < .001), Fig. 1f. Note that self-reported SRS T-scores did not correlate with MD or SWI (Fig. S4).

### Functional connectivity predicts longitudinal changes in adaptive function

From the sample of 150 autistic people, we collected ABC score data at T1 from 122 people, at T2 from 90 people, and at both timepoints from 81 people, Table 2.

**Table 2.** Vineland Adaptive Behaviour Composite (ABC) Scores by timepoint and age-bracket. T1 = timepoint 1, T2 = timepoint 2

|  | 6-14 years |  | 15-24 years |  | 25-31 years |  | 6-31 years |  |
| --- | --- | --- | --- | --- | --- | --- | --- | --- |
| Mean ABC T1 (SD) | 77.9 (9.9) | n=48 | 68.3 (13.1) | n=63 | 79.1 (22.0) | n=11 | 73.0 (13.8) | n=122 |
| Range | 58 – 100 |  | 30 – 104 |  | 52 – 121 |  | 30 – 121 |  |
| Mean ABC T2 (SD) | 77.2 (10.4) | n=46 | 72.8 (12.9) | n=41 | 81.3 (1.2) | n=3 | 75.3 (11.6) | n=90 |
| Range | 59 – 102 |  | 20 – 95 |  | 80 – 82 |  | 20 – 102 |  |
| Mean ABC change (SD) | -1.6 (9.0) | n=40 | 3.4 (10.8) | n=38 | 4.0 (10.8) | n=3 | 0.95 (10.1) | n=81 |
| Range | -23 - +26 |  | -37 - +30 |  | -5 - +16 |  | -37 - +30 |  |
| Mean follow-up interval (SD) | 18.9 (3.9) months |  | 20.7 (2.3) months |  | 18.0 (1.4) months |  | 19.7 (3.3) months |  |

#### Cross-sectional analyses between functional connectivity and adaptive function

First, we examined if MD and SWI were associated with T1 adaptive function abilities in autistic people. Lower MD and higher SWI were associated with lower T1 ABC across 6-31-year-olds (Tables S3-S4).

#### Longitudinal analyses between functional connectivity and adaptive function

Next, we assessed the association between T1 functional connectivity metrics and longitudinal changes in adaptive function. The mean follow-up interval between T1 and T2 ABC measurements across all autistic participants was 19.7 months (SD = 3.5 months), Table 2, Fig. S5a. ΔABC ranged from -37 to +30, Table 2, Fig. S5b. The 25-31-year-old group only had 3 participants with longitudinal data and were excluded from age-bin analyses. The mean follow-up interval was 1.8 months (SD = 3.9 months) longer in 15-24-year-olds compared to 6-14-year-olds, *t*_(68.5)_ = 2.3, *p* = .02. There were no differences in the variance of ΔABC between age-groups (F_2/78_ = .61, *p* = .55).

MD did not predict ΔABC, when 6-31-year-old autistic people were analysed together (Fig. 2a, Table S5). MD did not predict ΔABC in 6-14-year-old autistic people, Table S6.

**Figure 2.**
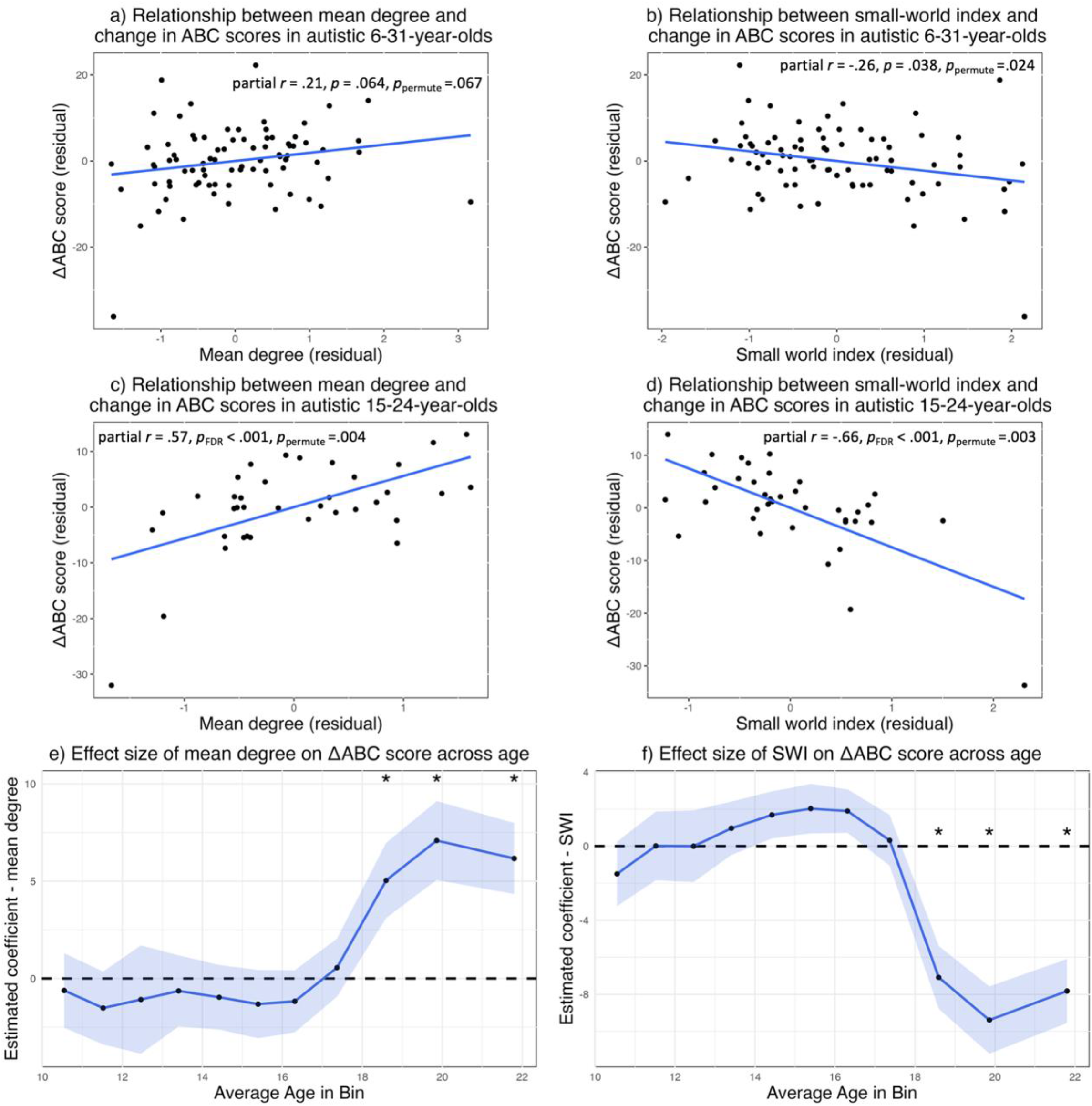
Mean degree and small-word index predict longitudinal changes in adaptive function. Relationships between timepoint 1 **a)** mean degree (MD), **b)** small-world index (SWI) and longitudinal changes in adaptive function in autistic people aged 6-31-years (n = 81). Predictive ability was stronger in autistic youth (15-24-years-old, n = 38), where adding the variable **c)** MD to the model predicting change in adaptive behaviour composite (ΔABC) score increased the amount of explained variance by 21%, while **d)** adding the variable SWI increased the amount of explained variance by 30%. **e-f)** We plotted a moving estimated coefficient of the functional connectivity metric predicting ΔABC as a function of average age, *without* pre-defined age-brackets. There were main effects of MD and SWI in models predicting ΔABC when the mean age in participant bins was between 18.6 to 21.8 years. *: *p*_FDR_ < .05. Blue ribbon = standard error.

In 15-24-year-old autistic people, higher MD was associated with improvements in ABC, with a large effect size (β = 5.6, SE = 1.5, *p*_FDR_ < .001, *p*_permute_ = .004, η^2^ = 0.13), Fig. 2c, Table S7. Adding the variable MD to a model predicting ΔABC (already containing full-scale IQ, T1 ABC, age and sex) increased the variance explained in ΔABC by 21% (model R^2^ increased from .29 to .50).

SWI predicted ΔABC when 6-31-year-old autistic people were analysed together (β = -2.4, SE = 1.0, *p* = .023, *p*_permute_ = .024, η^2^ = 0.04), Fig. 2b, Table S8. Adding the variable SWI to a model predicting ΔABC (already containing full-scale IQ, T1 ABC and age) increased the variance explained in ΔABC by 4% (model R^2^ increased from .30 to .34).

SWI did not predict ΔABC in 6-14-year-old autistic people, Table S9.

In 15-24-year-old autistic people, lower SWI was associated with improved ABC, with a large effect size (β = -7.5, SE = 1.6, *p*_FDR_ < .001, *p*_permute_ = .003, η^2^ = 0.18), Fig. 2d, Table S10. Adding the variable SWI to a model predicting ΔABC (already containing full-scale IQ, T1 ABC, age and sex) increased the variance explained in ΔABC by 30% (model R^2^ increased from .29 to .59).

Associations between actual and predicted values of ΔABC for the three models that significantly predicted ΔABC are shown in Figure S6. We assessed the performance of these models on ‘unseen’ participants using 10-fold cross-validation. The cross validation R^2^s were between .39 and .62 (Table S11).

A plausible trivial explanation for different effect sizes between the two age-brackets could be a between-age-bracket difference in the variances of SWI, MD or ΔABC. A *post hoc* Levene’s test showed that this was not the case (ΔABC: F_1/76_ = 0.20, *p* = .66; SWI: F_1/76_ = 1.4, *p* = .23; MD: F_1/76_ = .50, *p* = .48). Additionally, we examined *post hoc* if the strong association between functional connectivity and ΔABC in 15-24-year-olds was specific to ΔABC, or if it was also found in other brain-behaviour associations. Associations between SRS T-score and SWI/MD were stronger in younger participants (Figure S7).

As all age-bracket definitions are somewhat arbitrary, we illustrated the relationship between prediction ability for ΔABC and age *without* pre-defined age-brackets. We plotted a moving estimated coefficient of the functional connectivity metric predicting ΔABC as a function of average age. There were main effects of MD and SWI in models predicting ΔABC when the mean age in participant bins was between 18.6 (range 15.2-23.0) to 21.8 (range 17.5-30.3) years, the latter being the age-range ceiling in our sample, Fig. 2e-f, Table S12. This is consistent with better prediction ability in autistic youth compared to autistic children.

### Comparison against other prognostic variables

We compared the predictive performance of MD and SWI to that of autistic features and full-scale IQ. For the models with a significant main effect of functional connectivity metric on adaptive function outcomes, MD and SWI outperformed both behavioural predictors (Supplementary Results and Table S13).

### Categorical prediction of adaptive function changes

We explored the ability of the three significant regression models to categorise ΔABC (into improved, worsened and static outcomes) on a per-participant basis, using leave-one-out cross-validation. The categorisation accuracy of MD in 15-24-year-olds was 63.2%, *p*_permute_ = .002; SWI in 15-24-year-olds was 65.8%, *p*_permute_ = .001; SWI in 6-31-year-olds was 60.4%, *p*_permute_ < .001 (Fig. 3a). Thus, all models categorised ΔABC significantly better than chance. Then, we explored model performance in performing binary categorisation into increased (≥ 6) or not-increased (< 6) ΔABC. For the model containing MD in 15-24-year-olds, AUC = 0.84 [95% CI: 0.71-0.97], sensitivity = 0.73, specificity = 0.74; for SWI in 15-24-year-olds, AUC = 0.78 [95% CI: 0.63-0.93]; sensitivity = 0.73; specificity = 0.78; for SWI in 6-31-year-olds, AUC = 0.76 [95% CI: 0.63-0.89]; sensitivity = 0.43; specificity = 0.93 (Fig. 3b).

**Figure 3.**
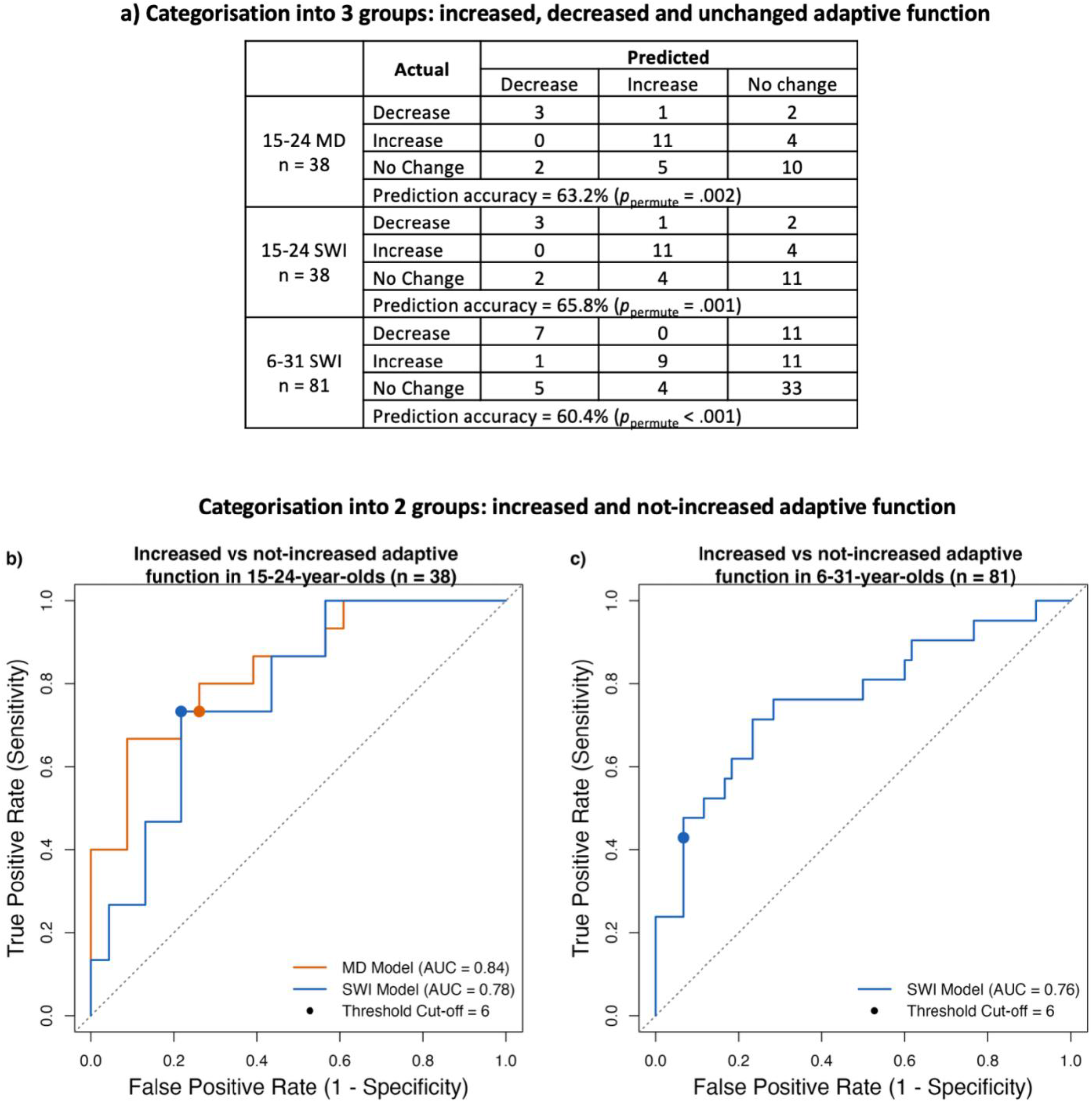
Illustrating the ability of models containing mean degree and small-world index to predict categorical changes in adaptive function based on minimal-clinically important difference thresholds, using leave-one-out cross-validation. Category thresholds were defined using a minimal-clinically important difference value of ≥ 6. **a)** All included models categorised adaptive function score outcomes into three categories significantly better than chance, as per permutation analyses. In **b)** 15-24-year-olds and **c)** 6-31-year-olds, receiver operator characteristic curves demonstrated good performance in binary categorisation into improvers and non-improvers. MD = mean degree; SWI = small-world index; AUC = area under the curve.

### Properties desired in candidate prognostic biomarkers

MD and SWI had good test-retest reliability (n = 203), both when derived from one epoch and from the mean of 2 epochs. For MD, ICC (2,1) = 0.78 [95% CI: 0.72-0.82] and ICC (2,k) = 0.88 [95% CI: 0.84-0.91]. For SWI, ICC (2,1) = .77 [95% CI: 0.71-0.82] and ICC (2,k) = 0.87 [95% CI: 0.83-0.90]. In the three models that significantly predicted ΔABC in autistic people, we performed a sensitivity analysis using MD/SWI derived from only one (instead of the mean of one-to-four) 20-second EEG epoch per participant, Fig. S8. The associations between MD/SWI and ΔABC were similar when only one epoch was used, although expectedly smaller in magnitude – consistent with the ICC values.

To assess for convergence with biological processes, we investigated if MD and SWI were associated with polygenic variation in autism- and brain-volume-related genes. We found no associations between polygenic scores (PGS) for autism and MD/SWI, Tables S14-S15.

PGS for brain volume was associated with variance in MD (n = 136, partial *r^2^* = .073, *p*_FDR_ = .004) and SWI (n = 136, partial *r^2^*= .090, *p*_FDR_ = .002), even when adjusting for age and sex, Fig. 4. Notably, no association between PGS for brain volume and MD/SWI was found in the non-autistic group, despite similar variances in PGS and MD/SWI between groups, Table S16.

**Figure 4.**
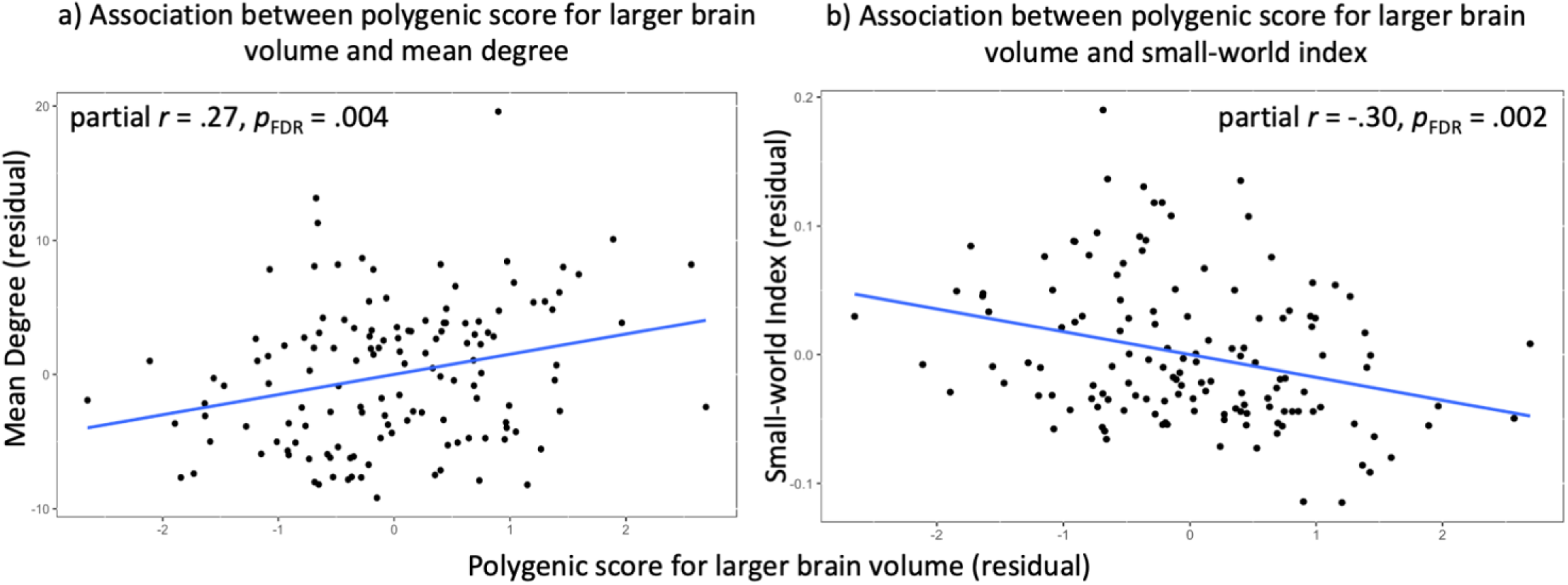
Relationship between polygenic score for brain volume and functional connectivity metrics. Polygenic scores for brain volume were significantly associated with variance in mean degree (partial *r^2^* = .073) and SWI (partial *r^2^* = .090), n = 136. A more positive polygenic score is associated with more alleles for large brain volume. These gene-brain relationships were only found in autistic people.

Given the association between PGS for brain volume and functional connectivity metrics, we confirmed *post hoc* that ΔABC was not trivially associated with head circumference (a proxy of brain volume; 98), Tables S17-S19.

## Discussion

We provide the first evidence that EEG-derived functional connectivity metrics can predict longitudinal changes in adaptive function in autistic people. By explicitly considering age-effects in a large, multi-site cohort, we demonstrated that mean degree and small-world index explain 21-30% of additional variance in outcomes in autistic youth (aged 15-24-years). SWI also significantly predicted adaptive function outcomes across autistic people aged 6-31 years, although it only explained 4% of additional variance in this age-bracket. Both metrics showed properties desired in prognostic biomarkers: convergence with underlying neurobiology (polygenic variation in brain volume) and high test-retest reliability.

Conflicting findings from previous autism functional connectivity studies has limited the development of functional connectivity metrics as biomarkers. Our cross-sectional analyses suggest that considering developmental stage in autism functional connectivity analyses may reconcile some of this heterogeneity, as has also been argued by Uddin *et al.* (40).

### Utility

In 15-24-year-old autistic people and 6-31-year-old autistic people (for SWI only), we found that EEG-derived MD/SWI were better predictors of changes in adaptive function than were behavioural predictors: autistic features and/or full-scale IQ. In 15-24-year-olds, our metrics predicted greater variance in changes in adaptive function (21-30%) than the N170 (14) or cortical structure (20), which both explained 6% of additional variance. Our study expands on an important but small (n = 27) fMRI functional connectivity study in a homogenous sample of autistic males, which also predicted similar variance in outcomes (17).

In categorising outcomes as increased, static and decreased on a per-person basis, based on minimal-clinically important differences (93,94) in ABC scores, models containing MD and SWI performed significantly better than chance, even when including participants across the 6-31-year age-range. Further, in binary categorisation into “increase” versus “no-increase” groups, the model containing MD in 15-24-year-old autistic people had an AUC of 0.84 and the model containing SWI in 6-31-year-old autistic people had an AUC of 0.76, suggesting promise as prognostic biomarkers (99).

Together, these findings suggest that EEG-derived MD and SWI may be developed as prognostic biomarkers for both continuous and categorical changes in adaptive function, especially for autistic people aged 15-24-years. In clinical contexts, this may help identify groups at risk of declining adaptive function during a key transitional period of development – enabling early, targeted interventions of adaptive functioning (e.g. the TEACCH School Transition to Employment and Postsecondary Education Program; 100). In clinical trials aiming to improve adaptive function (10,11,101), these biomarkers may be used to stratify participants by their likely spontaneous outcomes. This could reduce trial costs and sample size requirements (14).

### Brain-behaviour associations in different age groups

We identified the age-bracket of 15-24-years as that where MD and SWI are particularly effective predictors of adaptive function outcomes. The question is, *why*? Firstly, during youth, precision in the development of functional connectivity could be particularly important for behavioural outcomes. Therefore, variation in functional connectivity between youth may have a large effect on outcomes. Consistently, cortical and subcortical functional connectivity organisation (102) – including that involved in social cognition (67,68,103) – is significantly disrupted and remodelled between the ages of 14-26 years (67,68). In other age-groups, conservative maturation (where strong connections become stronger and weak connections become weaker) may be more pervasive (67,102); therefore, variation across individuals may have a less profound impact. Secondly, functional connectivity derived MD and SWI may be key drivers of adaptive function outcomes in late adolescence, whereas other (e.g. behavioural) features could be more important at other ages. Thirdly, the effect size of MD differences between autistic and non-autistic people was greatest in the 15-24-year age-group. That connectivity is most different from non-autistic people in this age-group may also suggest that it is the most relevant.

Additionally, we consider two explanations that might arise due to peculiarities of our dataset. It could be that brain-behaviour associations *in general* are strongest in the 15–24-year age-group. If true, associations between MD/SWI and other behavioural features (e.g. SRS T-scores) would also be higher in this age-group than others. We found that this was not the case. Next, it could be that the variances in ΔABC, MD or SWI between age-groups were different, allowing associations to be found in some but not other groups. This was also not found to be the case.

### The direction of relationships between functional connectivity and phenotype

MD was lower in autistic than non-autistic youth, in keeping with prior literature (40). Lower MD was associated with more autistic features and worsening of age-normed adaptive function over time. Regarding SWI, previous studies using heterogeneous methods and age-groups found both decreased (30) and increased (31) small-world network organisation in autistic compared to non-autistic children. We found that greater SWI was associated with more autistic features and worsening of age-normed adaptive function over time. However, network models postulate that higher SWI reflects an optimum balance between network integration and segregation (34). What may explain this apparent contradiction? Studies in other brain-based conditions have shown that less small-world organisation is associated with more favourable phenotypes (104,105). This may suggest that non-optimal connectivity *between* nodes may be protective in the context of altered function *within* nodes. Further, the argument that higher SWI equates to ‘better’ network organisation assumes constant MD (36); this was not the case across participants in our study (see limitations).

### Genetic associations with MD and SWI

Polygenic scores for brain volume (90) explained a high proportion of variance (in the context of polygenic scores; 14,106) in MD and SWI in autistic people. Whole brain, white matter and cortical volume differences have been repeatedly found in autistic people (20,43,44). Twin studies have shown that the heritability of MD is approximately 37% (107), while SWI has a heritability of approximately 50% (51,52). Our findings suggest that the effects of genotype on MD and SWI could explained by polygenic variation in genes contributing to brain volume.

Interestingly, the associations between PGS for brain volume and MD/SWI were only found in autistic people. This may suggest that genetic influences on brain volume affect MD and SWI more strongly in the co-presence of genetic and environmental factors that autistic people (and not non-autistic people) are exposed to. It is plausible that genetic variants for brain volume and autistic features interact in complex ways.

### Study strengths and limitations

Our study design had many strengths. We included autistic people at 5 study sites with co-occurring developmental and mental health conditions – reflecting autistic people in the general population (64). Thus, our findings may generalise to a large group of autistic people. Second, we used the Vineland Adaptive Behaviour Scale (69,108) – shown to have high test-retest reliability (69), convergent validity and internal consistency (109) – to measure adaptive function. Third, EEG is a widely available and affordable measure of functional connectivity (55), further improving translation potential.

We showed the robustness of MD/SWI as prognostic biomarkers. MD showed sensitivity to diagnostic group and both MD and SWI varied with the extent of parent-reported autistic features. Further, MD/SWI were associated with *current* adaptive function in the same direction as they were associated with changes in adaptive function. The models predicting longitudinal changes in adaptive function explained a high proportion of variance in outcome even in ‘unseen’ autistic participants, as shown by 10-fold-cross-validation. Finally, we demonstrated that strong predictive performance in autistic youth was not dependent on arbitrary age-bin definitions, finding significant predictive ability of MD/SWI across several age-bins that included autistic youth.

Our study has important limitations. Although our overall sample was large, we had a modest sample size of autistic youth with longitudinal Vineland data (n = 38). Further, we had few participants in the adult group (n = 32); therefore, group-difference analyses for MD/SWI in this age-group are likely underpowered. Next, we used the Humphries (85) method of quantifying SWI, which is affected by network density (MD; 36). Networks with higher MD had lower SWI (Fig. S9). Therefore, the two network metrics used are not independent. That said, this is true for most network metrics (110,111). Previous research has addressed this issue by thresholding network density across participants (112). However, this has the effect of excluding weaker functional connections. We decided against this approach as weak functional connections play an important role in cognition (113,114).

As expected, participants with more usable EEG epochs had a higher mean age. Therefore, the functional connectivity metrics, which were averaged across epochs, were more reliable in older participants (87). This is reassuring for our findings in autistic youth, but our negative findings in autistic children should be interpreted with some caution.

It is plausible that the ABC is measuring different adaptive behaviour constructs at different ages. For instance, the relationship between ABC and intelligence is stronger in children than in adults (18). If this is the case, our differential findings between children and youth could represent differences is *measurement* of adaptive behaviour rather than differences in brain-behaviour associations. Further work is needed to clarify the relevance of this limitation of the Vineland.

Studies of MCID in the ABC score (93,94) have predominantly included children; therefore, generalising MCID values to autistic adults should be done cautiously. Here, we used a relatively conservative MCID value of 6, consistent with the reliable change index of the ABC score.

We used chronological age as a proxy of developmental status. Developmental stage could have been captured more accurately through physiological-based measures such as Tanner (pubertal) staging; however, this would have also increased participant burden.

### Future work

Adaptive function skills are the amalgamation of many cognitive functions and their interactions with the environment. Future work could explore the cognitive abilities that mediate the effect of MD and SWI on longitudinal changes in adaptive function in autistic people. For example, recent findings from our group suggest that the organisation of EEG-derived large-scale functional connectivity networks is associated with complex facial emotion recognition in an age-sensitive manner (115). Similarly, future work should investigate if associations between MD/SWI and ΔABC are driven by certain adaptive function subdomains (i.e. communication, daily living or socialisation skills).

Most autistic people have a co-occurring mental health or neurodevelopmental condition (116), known to independently impact adaptive function (117). Future work could investigate relationships between co-occurring conditions, functional connectivity and longitudinal changes in adaptive function. Our sensitivity analysis without participants with ADHD, the most frequent co-occurring condition in our sample, found that relationships between MD/SWI and ΔABC remained consistent (Fig. S10).

The next steps in developing MD and SWI as prognostic biomarkers of adaptive function include assessing its performance in external, larger datasets (46). If such statistical validation is achieved, a randomised control trial is needed to investigate if prognostication improves the quality of life and/or employment, education and independent-living outcomes for autistic youth.

## Conclusion

Our study is the first to demonstrate that EEG-derived functional connectivity can explain a significant proportion of the variance in longitudinal changes in adaptive function in autistic people. Therefore, EEG-derived mean degree and small-world index may be developed as prognostic biomarkers for adaptive function outcomes, especially for autistic youth. Potential precision-medicine applications include stratifying autistic people in clinical trials and identifying those at risk of declining adaptive function in clinical settings. Finally, our age-sensitive approach may reconcile the heterogenous findings in previous autism functional connectivity studies.

## Data availability

The LEAP Study data for timepoints 1 and 2 is available here: https://www.aims-2-trials.eu/our-research/datasharing/

## Data Availability

The LEAP Study data is available here, subject to an approved data application: https://redcap.pasteur.fr/surveys/?s=YRFF78PH89

## Acknowledgments

We are grateful to the AIMS-2-TRIALS LEAP-group for data collection and quality control procedures. We are grateful to Dr Piotr J. Franaszczuk for his expert advice on signal analysis and filtering. We are grateful to the reviewers for their help in improving our study.

## Author Contributions

Chirag Mehra – Conceptualisation, Methodology, Formal analysis, Validation, Writing - Original Draft, Writing – Review & Editing, Visualization

Petroula Laiou – Resources, Software, Writing - Review & Editing

Pilar Garces - Data Curation, Formal analysis, Writing - Review & Editing

Joshua B Ewen – Writing - Review & Editing, Supervision

Eva Loth – Funding acquisition, Project administration

Mark H Johnson – Project administration, Funding acquisition

Luke Mason - Software, Project administration

Emily JH Jones – Writing - Review & Editing, Funding acquisition, Project administration

Tony Charman – Writing - Review & Editing, Funding acquisition

Thomas Bourgeron – Resources, Investigation, Funding acquisition

Jan Buitelaar – Writing - Review & Editing, Funding acquisition, Project administration

Michael Absoud - Writing - Review & Editing, Supervision

Mark P Richardson – Supervision, Funding acquisition

Declan Murphy – Conceptualisation, Funding acquisition, Project administration

Jonathan O’Muircheartaigh - Writing - Review & Editing, Supervision, Funding acquisition

## Funding

This work was supported by EU-AIMS (European Autism Interventions), which received support from the Innovative Medicines Initiative Joint Undertaking under grant agreement no. 115300, the resources of which are composed of financial contributions from the European Union’s Seventh Framework Programme (grant FP7/2007-2013), from the European Federation of Pharmaceutical Industries and Associations companies’ in-kind contributions, and from Autism Speaks. The results leading to this publication have also received funding from the Innovative Medicines Initiative 2 Joint Undertaking under grant agreement No 777394 for the project AIMS-2-TRIALS. This Joint Undertaking receives support from the European Union’s Horizon 2020 research and innovation programme and EFPIA and AUTISM SPEAKS, Autistica, and SFARI. This study was also delivered through the National Institute for Health and Care Research (NIHR) Maudsley Biomedical Research Centre (BRC). JOM was funded by a Sir Henry Dale Fellowship jointly by the Wellcome Trust and the Royal Society (206675/Z/17/Z). JOM received support from the Medical Research Council Centre for Neurodevelopmental Disorders, King’s College London [MR/N026063/1]. None of the funders had a role in the design of the study; in the collection, analyses, or interpretation of data; in the writing of the manuscript, or in the decision to publish the results. Any views expressed are those of the author(s) and not necessarily those of any of the funders, the NIHR or the Department of Health and Social Care.

## Conflicts of interest

Joshua B Ewen previously consulted for Novartis. In the past 3 years, Jan K Buitelaar has been a consultant to / member of advisory board of / and/or speaker for Takeda, Medice, Angelini, Neuraxpharm and Bitsphi. He is not an employee of any of these companies, and not a stock shareholder of any of these companies. He has no other financial or material support, including expert testimony, patents, royalties. Pilar Garcés is employed by Roche Innovation Center, Basel, Switzerland. Tony Charman has received consultancy fees from F. Hoffmann-La Roche Ltd. and royalties from Sage Publishing and Guilford Press. No other author has competing interests to declare.

## Supplementary material

Supplementary material is available online.

## Notes

### Author Declarations

London - Queen Square Research Ethics Committee gave ethical approval for this work

### Summary of Updates

Updated analyses, results, figures, limitations and discussion.

