## Supplemental File for "EEG functional connectivity as a prognostic biomarker of adaptive function in autistic people"

Supplementary materials

### Supplementary Methods

**LEAP Study sites**

Participants in our analyses were from 5 study sites of the AIMS-2-TRIALS Longitudinal European Autism Project (LEAP): the Central Institute of Mental Health (CIMH, Mannheim, Germany), King’s College London (KCL, United Kingdom), Radboud University Medical Centre (RUMC, Netherlands), University Campus BioMedico (UCB, Rome, Italy) and University Medical Centre Utrecht (UMCU, Netherlands).

At each site, participants were recruited from a variety of sources including existing volunteer databases, existing research cohorts, clinical referrals from local outpatient centres, special needs schools, mainstream schools and/or local communities. Recruitment pathways and strategies were optimised to each site.

**Participant co-occurring conditions**

We ascertained diagnoses of co-occurring mental health, developmental, neurological and other conditions through use of both a parent- (for child participants) or self-reported (for adult participants) survey and a structured Family Medical History Interview (1).

**Behavioural measures**

The age-standardised ‘T-scores’ of the Social Responsiveness Scale-Second Edition (SRS-2; 2) were used as trait measures of autistic behaviours. The SRS-2 T-score is highly reliable (2). There was no significant association between self- and parent- reported SRS T-score in autistic people, Figure S1. We used the parent-reported instead of self-reported T-scores in our main analysis, as we had a larger sample size with the former (n = 118 versus n = 76), and the values are not interchangeable (Fig. S1).

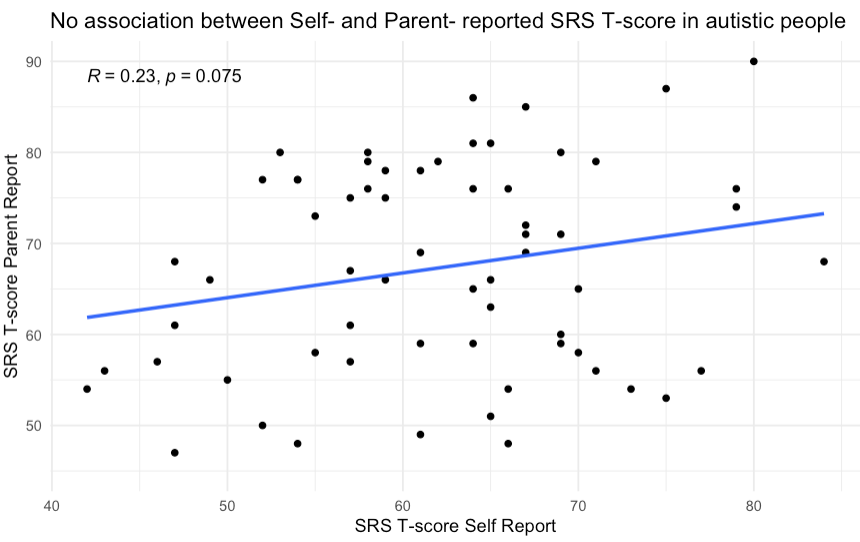

**Figure S1 –** There was no significant association between self- and parent- reported SRS T-score in autistic people.

Intellectual ability was assessed using the Wechsler Abbreviated Scales of Intelligence – Second Edition (3), or the shortened versions of the German, Dutch or Italian Wechsler Intelligence Scale for Children, third or fourth editions (4,5) or Wechsler Adult Intelligence Scale third or fourth editions (6,7; as the WASI-II is not translated into these languages). The shortened versions were used to minimise testing burden on participants. All used measures included two verbal subscales (Vocabulary, Similarities) and two non-verbal subscales (Block Design, Matrix Reasoning). These measures of full-scale IQ from the shortened scales are highly correlated (*r* = .93) with a full-scale IQ obtained by administering the complete test. Age-standardised estimates of intellectual functioning were derived from national population norms.

**EEG data acquisition**

Electrodes were arranged in a 10-20 layout. The following EEG signal acquisition systems were employed: Brainvision (CIMH, KCL, RUNMC), Biosemi (UMCU) and Micromed (UCBM), with sampling frequencies of 5000 Hz (KCL, RUNMC), 2048 Hz (UMCU), 2000 Hz (CIMH) and 256–1000 Hz (UCBM).

EEG data processing and analyses were performed using MATLAB and custom scripts.

**EEG Processing**: We treated the whole 20-second epoch as a single window. Producing multiple windows has the advantage of better capturing moment-to-moment connectivity. However, longer window-length increases the association between the functional connectivity strength quantified empirically and the ‘true’ functional connectivity strength, as per simulations (8).

Prior to source localisation, initial downsampling and bandpass filtering were performed, as per Hyvarinen (9). Except for signals from UCBM (which were recorded at 256 Hz and not initially downsampled), EEG signals were downsampled to 1000 Hz, with an antialiasing filter (low-pass finite impulse response filter, Kaiser window, cutoff of 300 Hz, transition band width 100 Hz), using EEGlab (10). Then, the entire EEG resting-state recording (all four 30 second epochs and the segments in between them), with 10 seconds of data padding, was bandpass filtered between 1–32 Hz with a finite impulse response filter of order 2000, using a Hamming window, in both forward and backward directions, with detrending, using fieldtrip. This relatively high filter order (11) was used as high pass filtering was performed for frequencies less than 1 Hz and to strongly penalise muscle artefact (> 32 Hz).

**EEG artefact cleaning**

EEG artefact identification and removal was performed for each 30 second epoch, as per Garcés *et al.* (12). Only data from the 61 electrodes commonly used across all sites were retained. The following steps were performed: (1) electrode channels with poor data quality were removed, (2) temporal segments with large transient artifacts such as muscle bursts or movements were discarded, (3) independent component analyses were applied using fastICA(9) to distinguish signal from noise, (4) artefactual independent components were identified and eliminated, (5) and channels discarded in (1) were interpolated. The common average signal across all channels was used as the reference. If more than 10 cannels were eliminated in step 1, the epoch was not included in subsequent analyses.

**Source Reconstruction**

Source reconstruction was performed using individual-participant T1-weighted MRI images. T1-weighted MRIs were segmented with Statistical Parametric Mapping 12 (SPM12) software (13) into grey matter, white matter, cerebrospinal fluid, bone, soft tissue and air. These probabilistic images were then smoothed (5mm full width half maximum), thresholded and resliced to produce binary masks of 2mm × 2mm × 2mm resolution for three tissue types: brain (including grey matter, white matter and cerebrospinal fluid), skull and scalp. These binary masks were transformed to hexahedral meshes with FieldTrip (14). All three considered tissue types were assumed to have homogeneous and isotropic conductivity: 330 ms/m for the brain and scalp (15), and an age-dependent skull conductivity of 3.958  +  62.77 x *e*^− 0.2404 x age in years^ ms/m, in line with the BESA (16) recommended conductivity ratios. Segmentations were visually inspected. The forward model was derived with FieldTrip and SimBio (17). For the inverse model, 1200 source locations of interest were defined in grey matter in Montreal Neurological Institute (MNI) space, following a 3D cubic diamond grid. Source positions were transformed from MNI space to each subject’s individual space with a nonlinear transformation using SPM12. Electrode positions were determined by transforming standard MNI positions to subject-space with the same transformation, then projecting to the scalp surface. Source time series were estimated with linearly constrained minimum variance beamformer (18), using a regularization of 5% of the average trace of the covariance matrix. The 1200 sources were parcelled into 68 cortical regions of interest, as per the Desikan-Killiany atlas (19). Each region was comprised of between 1 and 67 sources (median of 13). Where there was more than one source per region, the representative time series for a given region was defined as the first principal component of all the source time series in it.

**Filtering**

First, we downsampled the signals to 250Hz and performed data quality checks. Then, we filtered signals between 6-9 Hz. High-pass followed by low-pass filters were used in sequence, using two-pass (using the MATLAB function filtfilt), finite impulse response filters, of order 250. To reduce signal distortion, data padding of 500 samples was used during filtering and while performing Hilbert transformations.

We filtered signals between 6-9 Hz (‘low-alpha’) for the following reasons: first, small-world index based on 6-9 Hz EEG signals has been found to have the highest test-retest reliability among frequency bands between 0.5-45 Hz (20). Second, functional connectivity differences between autistic and non-autistic people have been found in the theta and alpha frequency bands (21). While the traditional limits of these bands are arbitrarily defined (22), statistical factor analyses of EEG spectra (23) revealed that the data-driven factor with the widest frequency band between 4-13 Hz is 6-9Hz. Finally, the 6-9Hz band may be sensitive to developmental changes in neurobiology: a metanalysis (24) of spectral power changes through development found that peak alpha frequency shifted from 5.4 Hz in toddlerhood to 9.9 Hz in late adolescence.

**The** **phase locking vale (PLV)**

The PLV has the following advantages as an EEG functional connectivity method: it is largely independent of amplitude relationships between brain-regions; scalp EEG-derived PLV has been shown to have high test-retest reliability, concordance with its underlying structural connectivity and ability to predict longitudinal changes in behaviour (25). Importantly, the PLV is sensitive to age-related connectivity changes (25).

The PLV between the signals $x_{i}(t)$ and $x_{j}(t)$ , $t = 1,\ldots., N$ where $N$ is the number of points in an EEG epoch is defined as:

$${PLV}_{ij}= \frac{1}{N}\left| \sum_{t=1}^{N} e^{i*\Delta\theta(t)} \right|,$$

where $\Delta\theta(t)$ is the difference of the instantaneous phases $\theta_{i} (t),\theta_{j}(t)$of the signals $x_{i}(t), x_{j}(j)$. We used permutation testing to determine statistically significant edges.

**Permutation testing with the phase locking vale (PLV)**

An advantage of the PLV is that permutation testing (based on surrogate data) is innate to the method (26). This attempts to differentiate ‘true’ connectivity from connectivity that may appear due to noise. For each real timeseries from each brain-region, we created 99 surrogate timeseries, with matching amplitude spectrum and signal distribution as the real data (27). PLV values for each of the surrogate timeseries were calculated. PLV values between two regions in the real timeseries were only retained if the value was in the top 5% PLV values (real and surrogate); otherwise, the PLV value between these brain regions was assigned a value of 0. Hence, only connections with a < .05 probability of occurring due to noise were retained.

**Calculating the normalised weighted clustering coefficient and the normalised weighted pathlength**

We calculated the weighted clustering coefficient for each node, as per Onnela *et al.* (28):

$\tilde{C_{i}}=\frac{2}{k_{i}(k_{i}-1)}\sum_{j,k} \left( \tilde{w}_{ij}\tilde{w}_{jk}\tilde{w}_{ki} \right)^{1/3}$,

where $k_{i}$ is the degree of node $i$, $\tilde{w}$ is the weighted adjacency matrix and *j* and *k* are other nodes in the network. Note that the weights in the weighted matrix were scaled by the largest weight in each matrix:

$\tilde{w}_{ij}=w_{ij}/max(w_{ij})$,

where *w* is the (non-weighted) adjacency matrix. To calculate the normalised clustering coefficient, we used a normalisation process by creating 500 random networks by randomising the empirical network, while preserving the degree and strength distributions: each edge was rewired 5 times, with edge weights randomised every 5th step. The weighted clustering coefficient of each random network was calculated, and then averaged across each of the 500 random networks to produce $C_{rand}$. Thus, normalised, weighted clustering coefficient for each node was calculated as:

$$C_{i}'=\frac{\tilde{C_{i}}}{C_{rand}}$$

$C_{i}'$at each node was averaged across 68 nodes to produce the mean normalised, weighted clustering coefficient (further referred to as normalised, weighted clustering coefficient). The mean normalised, weighted clustering coefficient across all nodes quantifies the intensity of interconnectedness between neighbouring nodes in a network, reflecting its potential for segregated information processing.

Next, we calculated the weighted, normalised path length. First, network edges were inverted to lengths (edges with larger weights produce shorter lengths). Then, the *shortest possible* distance between all pairs of nodes was computed. Finally, the average shortest distance between all pair of nodes produced the weighted normalised path length. Formally, the shortest weighted path length $d_{ij}$ between any two nodes, e.g. $i$ and $j$ is defined as:

$d_{i,j}^{w}=\sum_{w_{ij}\in gi\leftrightarrow j} w_{ij}$,

where $gi\leftrightarrow j$ is the shortest weighted path between nodes $i$ and $j$.

The weighted, average shortest pathlength (*L*) between node $i$ and all other nodes of the network is defined as:

$L_{i}=\frac{\sum_{i\neq j} d_{i,j}^{w}}{(N-1)}$  ,

where $N$ is the number of nodes (68 in this study) and $d_{i,j}$ is the shortest path length between nodes $i$ and $j$, when $j$ is any node that is not $i$.

The normalisation procedure was undertaken using the same methods as for the normalised weighted clustering coefficient, producing the normalised average shortest pathlength of node $i$:

$$L_{i}'=\frac{L_{i}}{L_{rand}}$$

$L_{i}'$ at each node was averaged across 68 nodes to produce the mean normalised, weighted pathlength (further referred to as normalised, weighted pathlength). The normalised weighted pathlength is a measure of functional integration or efficiency of the network: how efficiently information from one node can be transmitted to another.

**Test-retest reliability**

We quantified the intrasession test-retest reliability between two epochs recorded between 1-3 minutes of each other. Intrasession test-retest reliability was used to reduce the confounding effects of circadian rhythm (29) and caffeine (30) on functional connectivity. Reliability was calculated for metrics derived from one 20-second epoch using interclass correlation coefficient (ICC (2,1)) and metrics derived from the mean of two 20-second epochs using ICC(2,k). ICC values were interpreted as per Ku and Li (31): 0.9 -1.0 = excellent reliability, 0.75-0.9 = good, 0.5-0.75 = moderate and 0.0-0.5 = poor reliability.

**Genetics**

We calculated polygenic likelihood scores (PGS) using blood samples. Single-nucleotide polymorphism genotyping was undertaken by the “Centre National de Recherche en Génomique Humaine” (Paris, France), using the Infinium OmniExpress-24v1 BeadChip (>700 K markers) from Illumina. Using genome-wide association study summary statistics as a reference, PGS were produced for autism (32) and brain volume (33). For brain volume PGS, a more positive value is associated with more alleles for large brain volume. PGS scores were adjusted for ancestry and the raw PGS for each participant for each trait was standardized (z-scored) using European non-autistic participants as a reference. PGS were available for 136 autistic people and 133 non-autistic people.

**Statistical analyses**

We used the False Discovery Rate to correct for multiple comparisons (35); one comparison family was comprised of one hypothesis. All reported *p* values are two-tailed, α = .05. Effect sizes of independent variables in general linear models were measured using Type II Eta squared (η^2^). η^2^ = 0.01 indicates a small effect, η^2^ = 0.06 indicates a medium effect, and η^2^ = 0.14 indicates a large effect (38).

We standardised (z-scored to sample mean and standard deviation) values of mean degree, small-world index, age, full-scale IQ and follow-up interval to allow comparisons between the coefficient estimates from different general linear models.

General linear model assumptions (e.g. normality of residuals and homoskedasticity) were tested using standard approaches. For the assumption of linearity, the Ramsey Regression Equation Specification Error Test (39) was used to assess whether the addition of non-linear combinations of the independent variables improved model fit. This was limited to altering the age variable to its quadratic form, only, for reasons of interpretability.

We evaluated the out-of-sample generalisability of the models predicting longitudinal changes in ABC score using the connectome-based predictive modelling protocol (40). First, we plotted the predicted values from the model against the actual values from the data and fitted a regression line to assess bias (Figure S6). The gradient of this line was interpreted as follows: gradient ~ 1: predicted values closely match observed values, gradient < 1: model underestimates true values, gradient > 1: model overestimates true values. Then, we performed 5-fold cross-validation to assess model performance on ‘unseen’ participants. Finally, we tested the significance of the independent variables mean degree and small-world index – when adjusting for the presence of co-variates – using permutation testing, using the double residualisation permutation method (robust in the presence of high nuisance co-variate:sample size ratio; 41). We randomly assigned the ABC score changes to different participants and derived the correlation coefficient (*r*) for actual versus predicted values 10,000 times.

We used a minimal clinically important difference (MCID) of the Vineland Adaptive Behaviour Composite (ABC) Score of 6, based on studies (42,43) that calculated MCID for autistic people without intellectual disability in large samples. These studies used distribution-based and anchor-based methods (the latter comprising of external validation with clinician-reported assessments of adaptive function) to calculate MCID. Thus, MCID considers both the reliability of the ABC *and* changes in the ABC that are minimally required to be noted by a clinician. An MCID cut-off of 6 is consistent with the reliable change index (RCI; 42,44,45) of the ABC in autistic people without an intellectual disability (RCI = 6.2). The RCI identifies the value at which a change in ABC is unlikely (*p* < .05) to occur by chance given its standard error of measurement.

Participants per age-bracket with longitudinal ABC data

**Figure S2** – Distribution of participant ages in those with longitudinal adaptive function composite (ABC) score data. Children (6-14-years), n = 40; youth (15-24-years), n = 38; adults (25-31-years), n = 3. Given the small number of adult-group participants, this group were excluded from age-bracket analyses. Green lines = age-group boundaries.

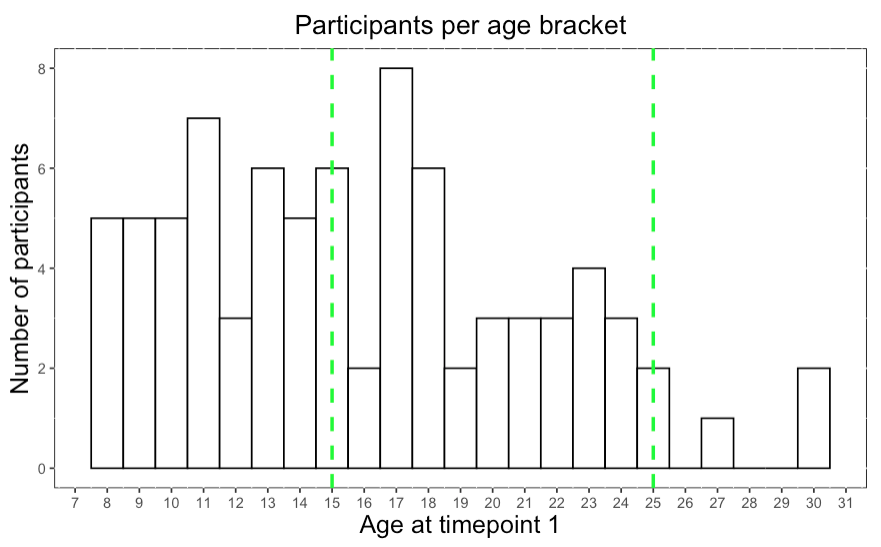

### Supplementary Results

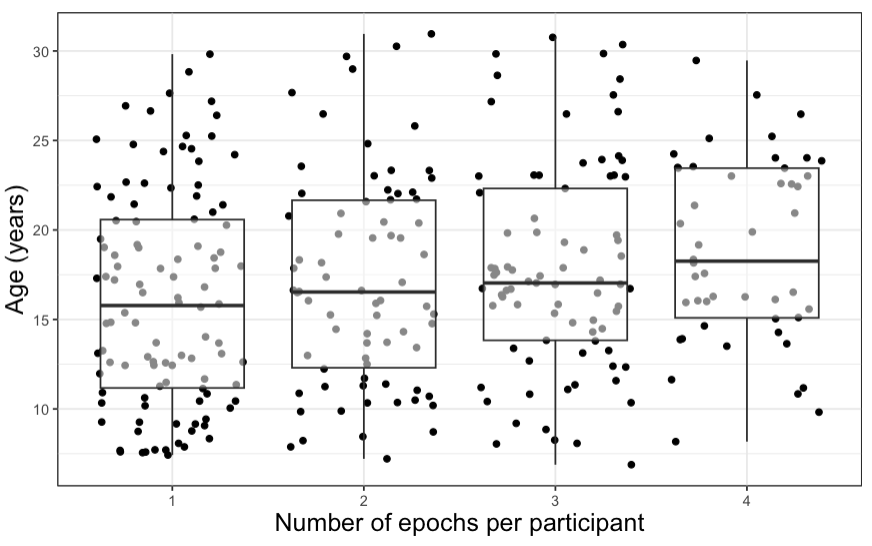

*H*(3) = 8.9, *p* = .03

**Figure S3** – Participants with more usable 20-second EEG epochs had a greater mean age

Relationship between number of epochs used to calculate mean degree or small-world index and participant age

**General linear models for group differences in mean degree and small-world index**

| **Table S1:** General linear model showing relationship between standardised mean degree, age and autism diagnostic status in 6-31-year-olds | | | | | |
| --- | --- | --- | --- | --- | --- |
| **N = 309** | **β** | **Std. Error** | **t** | ***p*** | **η^2^** |
| Intercept | 0.41 | 0.11 | 3.6 | < .001 | - |
| Group (autism) | -0.26 | 0.10 | -2.7 | .008 | 0.02 |
| Age | 0.64 | 0.07 | 9.0 | < .001 | 0.08 |
| Age^2^ | -0.18 | 0.05 | -3.9 | < .001 | 0.04 |
| Site 2 | -0.40 | 0.17 | -2.4 | .016 | 0.06 |
| Site 3 | 0.14 | 0.13 | 1.1 | .279 |  |
| Site 4 | -0.54 | 0.19 | -2.7 | .006 |  |
| Site 5 | -0.29 | 0.14 | -2.0 | .048 |  |
| **Group (autism) x Age** | **-0.19** | **0.10** | **-2.0** | **.045** | **0.01** |
| **Mean degree** ~ Group + Age + Group x Age + Age^2^ + site. R^2^ = .31, R^2^_adj_ = .29, F(8,300) = 17.02, p < .001. | | | | | |

| **Table S2:** General linear model showing relationship between standardised small-world index, age and autism diagnostic status in 6-31-year-olds | | | | | |
| --- | --- | --- | --- | --- | --- |
| **N = 309** | **β** | **Std. Error** | **t** | ***p*** | **η^2^** |
| Intercept | -0.23 | 0.12 | -1.9 | .061 | - |
| Group (autism) | 0.14 | 0.10 | 1.4 | .132 | 0.01 |
| Age | -0.58 | 0.08 | -7.6 | < .001 | 0.08 |
| Age2 | 0.19 | 0.05 | 3.9 | < .001 | 0.04 |
| Site 2 | 0.21 | 0.18 | 1.2 | .229 | 0.02 |
| Site 3 | -0.19 | 0.13 | -1.4 | .155 |  |
| Site 4 | 0.12 | 0.21 | 0.6 | .576 |  |
| Site 5 | -0.04 | 0.15 | -0.2 | .805 |  |
| **Group (autism) x Age** | **0.21** | **0.10** | **2.1** | **.039** | **0.01** |
| **Small-world index** ~ Group + Age + Group x Age + Age^2^ + Site. R^2^ = .23, R^2^_adj_ = .21, F(8,300) = 11.47, p < .001. | | | | | |

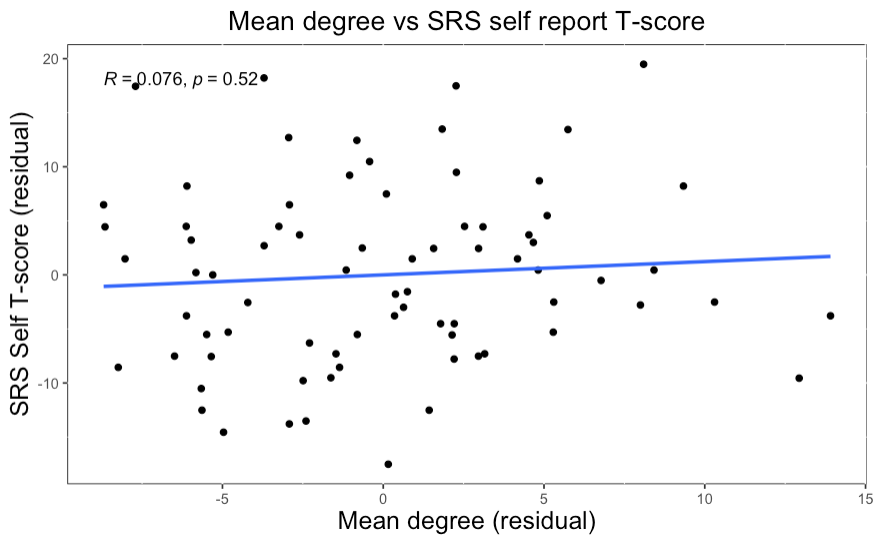

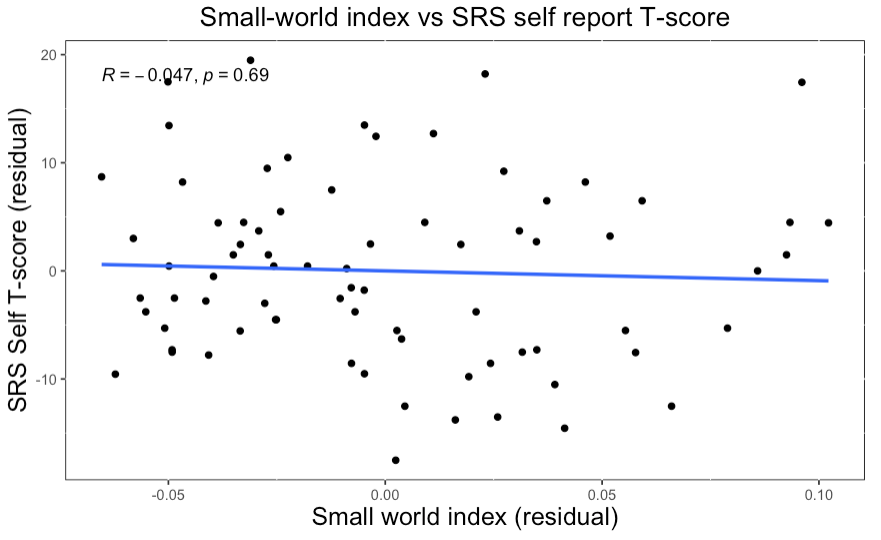

**Figure S4 –** Mean degree and small-world index were not significantly associated with self-report SRS T-scores.

**Cross-sectional analyses between functional connectivity and adaptive function**

| **Table S3:** General linear model showing relationship between mean degree, timepoint 1 ABC scores and age in autistic people aged 6-31 years | | | | | |
| --- | --- | --- | --- | --- | --- |
| **N = 122** | **Est. coeff.** | **Std. Error** | **t** | ***p*** | **η^2^** |
| Intercept | 120.00 | 11.06 | 10.85 | < .001 | - |
| **Mean degree** | **2.79** | **1.17** | **2.38** | **.019** | **0.03** |
| Sex (male) | -7.33 | 2.40 | -3.06 | .003 | 0.06 |
| Site 2 | 6.99 | 2.55 | 2.75 | .007 | 0.07 |
| Site 3 | 10.98 | 4.29 | 2.56 | .012 |  |
| Site 4 | 7.70 | 3.19 | 2.41 | .017 |  |
| Age | -4.89 | 1.25 | -3.93 | < .001 | 0.09 |
| Age^2^ | 0.11 | 0.03 | 3.34 | .001 | 0.07 |
| Timepoint 1 ABC score ~ mean degree + Age + Age^2^ + site + sex. R^2^ = .33, R^2^_adj_ = .28, F(7,114) = 7.8, *p* < .001. | | | | | |

| **Table S4:** General linear model showing relationship between small-world index, timepoint 1 ABC scores and age in autistic 6-31-year-olds | | | | | |
| --- | --- | --- | --- | --- | --- |
| **N = 122** | **Est. coeff.** | **Std. Error** | **t** | ***p*** | **η^2^** |
| Intercept | 121.27 | 11.18 | 10.84 | < .001 | - |
| **Small-world index** | **-2.84** | **1.16** | **-2.45** | **.016** | **0.04** |
| Sex (male) | -7.08 | 2.39 | -2.97 | .004 | 0.05 |
| Site 2 | 6.61 | 2.56 | 2.58 | .011 | 0.06 |
| Site 3 | 8.97 | 4.30 | 2.09 | .039 |  |
| Site 4 | 6.84 | 3.19 | 2.14 | .034 |  |
| Age | -5.10 | 1.27 | -4.03 | < .000 | 0.10 |
| Age^2^ | 0.12 | 0.03 | 3.51 | .001 | 0.07 |
| Timepoint 1 ABC score ~ small-world index + Age + Age^2^ + site + sex. R^2^ = .33, R^2^_adj_ = .28, F(7,114) = 7.8, *p* < .001. | | | | | |

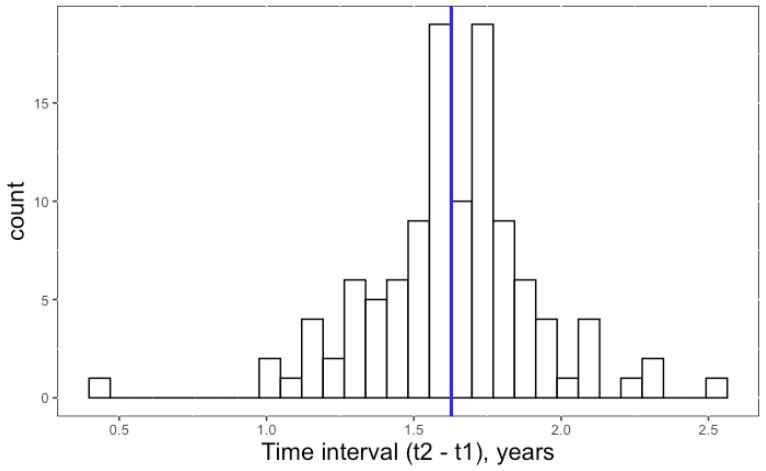

a

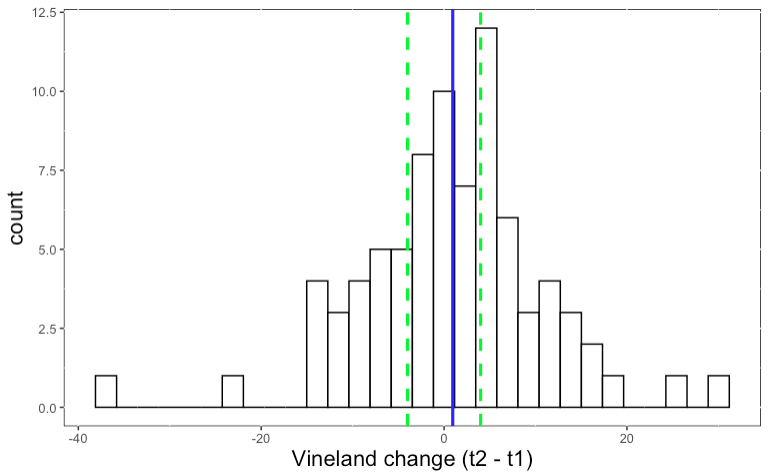

**Figure S5** – **a)** Time interval between timepoint 1 and timepoint 2 Vineland measurements, n = 81, mean = 1 year 7 months (SD = 3.5 months). **b)** Differences in Vineland composite score between timepoint 1 and 2. Mean = 0.95 (SD = 10.14). Blue lines show the means of each value. The green lines in **b)** show the minimal clinically important differences on the Vineland (i.e. the thresholds beyond which changes in Vineland composite score that are thought to be clinically significant).

b

**General linear models of mean degree predicting longitudinal changes in ABC scores**

| **Table S5:** General linear model showing relationship between mean degree, longitudinal changes in ABC scores, age and full-scale IQ in autistic 6-31-year-olds | | | | | |
| --- | --- | --- | --- | --- | --- |
| **N = 81 -** 6-31-year-olds | **Est. coeff.** | **Std. Error** | **t** | ***p*** | **η^2^** |
| Intercept | -1.8 | 2.0 | -0.9 | .355 |  |
| **MD** | **2.0** | **1.1** | **1.8** | **.077** | **0.03** |
| FSIQ | 1.1 | 0.9 | 1.2 | .232 | 0.01 |
| T1 ABC score | -8.6 | 1.9 | -4.6 | < .001 | 0.18 |
| Age | 0.9 | 1.2 | 0.7 | .464 | 0.01 |
| Follow-up interval | -5.6 | 1.4 | -3.9 | < .001 | 0.06 |
| Site 2 | 2.0 | 2.3 | 0.9 | .392 | 0.04 |
| Site 3 | 6.2 | 2.9 | 2.1 | .036 |  |
| Age x Follow-up interval | -4.1 | 1.4 | -3.0 | .004 | 0.07 |
| Longitudinal change in ABC score ~ MD + FSIQ + Timepoint 1 ABC score + Age + Follow-up interval + Site + Age x Follow-up interval. R^2^ = .39, R^2^_adj_ = .32, F(8,72) = 5.7, *p* < .001. | | | | | |

| **Table S6:** General linear model showing relationship between mean degree, longitudinal changes in ABC scores and age in autistic 6-14-year-olds | | | | | |
| --- | --- | --- | --- | --- | --- |
| **N = 40 -** 6-14-year-olds | **Est. coeff** | **Std. Error** | **t** | **p** | **η^2^** |
| Intercept | 0.6 | 4.1 | 0.1 | .892 |  |
| **MD** | **-1.3** | **1.6** | **-0.8** | **.404** | **0.02** |
| T1 ABC score | -2.6 | 2.1 | -1.3 | .213 | 0.04 |
| Age | 6.5 | 3.9 | 1.7 | .106 | 0.06 |
| Follow-up interval | -1.9 | 1.3 | -1.5 | .140 | 0.05 |
| Sex (male) | 3.8 | 3.0 | 1.3 | .217 | 0.04 |
| Site 2 | -0.5 | 3.6 | -0.2 | .880 | 0.05 |
| Site 3 | 4.5 | 3.8 | 1.2 | .249 |  |
| Longitudinal change in ABC score ~ MD + Timepoint 1 ABC score + Age + Follow-up interval + Site + Sex. R^2^ = .31, R^2^_adj_ = .15, F(7,32) = 2.0, *p* = .089. | | | | | |

| **Table S7:** General linear model showing relationship between mean degree, longitudinal changes in ABC scores, age and full-scale IQ in autistic 15-24-year-olds | | | | | |
| --- | --- | --- | --- | --- | --- |
| **N = 38 -** 15-24-year-olds | **Est. coeff.** | **Std. Error** | **t** | ***p*** | **η^2^** |
| Intercept | 4.2 | 3.7 | 1.1 | .271 |  |
| **MD** | **5.6** | **1.5** | **3.7** | **.001** | **0.13** |
| FSIQ | 4.5 | 1.5 | 3.0 | .005 | 0.09 |
| T1 ABC score | -8.8 | 1.7 | -5.2 | < .001 | 0.25 |
| Age | 0.0 | 0.0 | -1.1 | .282 | 0.05 |
| Follow-up interval | 22.1 | 13.8 | 1.6 | .121 | 0.1 |
| Sex (male) | -6.8 | 3.2 | -2.1 | .041 | 0.04 |
| Site 2 | 5.0 | 3.0 | 1.7 | .108 | 0.04 |
| Site 3 | 7.4 | 4.2 | 1.8 | .091 |  |
| Age x Follow-up interval | 0.0 | 0.0 | -2.1 | .047 | 0.04 |
| Longitudinal change in ABC score ~ MD + FSIQ + Timepoint 1 ABC score + Age + Follow-up interval + Site + Sex + Age x Follow-up interval. R^2^ = .62, R^2^_adj_ = .50, F(9,28) = 5.2, *p* < .001. | | | | | |
| Model statistics where MD is omitted as a variable: R^2^ = .44, R^2^_adj_ = .29, F(8,29) = 2.9, *p* = .017. Change in R^2^_adj_ on adding MD to model = +.21. | | | | | |

**General linear models of small world-index predicting longitudinal changes in ABC scores**

| **Table S8:** General linear model showing relationship between small-world index, longitudinal changes in ABC scores, age and full-scale IQ in autistic 6-31-year-olds | | | | | |
| --- | --- | --- | --- | --- | --- |
| **N = 81 -** 6-31-year-olds | **Est. coeff.** | **Std. Error** | **t** | ***p*** | **η^2^** |
| Intercept | -1.5 | 2.0 | -0.7 | .458 |  |
| **SWI** | **-2.4** | **1.0** | **-2.3** | **.023** | **0.04** |
| FSIQ | 1.0 | 0.9 | 1.2 | .245 | 0.01 |
| T1 ABC score | -8.4 | 1.8 | -4.6 | < .001 | 0.18 |
| Age | 1.2 | 1.1 | 1.0 | .305 | 0.01 |
| Follow-up interval | -5.6 | 1.4 | -4.0 | < .001 | 0.07 |
| Site 2 | 1.3 | 2.3 | 0.6 | .566 | 0.03 |
| Site 3 | 5.4 | 2.9 | 1.9 | .061 |  |
| Age x follow-up interval | -3.7 | 1.3 | -2.8 | .007 | 0.06 |
| Longitudinal change in ABC score ~ **SWI** + FSIQ + Timepoint 1 ABC score + Age + Follow-up interval + Site + Age x Follow-up interval. R^2^ = .40, R^2^_adj_ = .34, F(8,72) = 6.1, *p* < .001. | | | | | |
| Model statistics where SWI is omitted as a variable: R^2^ = .36, R^2^_adj_ = .30, F(7,73) = 5.8, *p* < .001. Change in R^2^_adj_ on adding SWI to model = +.04. | | | | | |

| **Table S9:** General linear model showing relationship between small-world index, longitudinal changes in ABC scores and age in autistic 6-14-year-olds | | | | | |
| --- | --- | --- | --- | --- | --- |
| **N = 40 -** 6-14-year-olds | **Est. coeff.** | **Std. Error** | **t** | ***p*** | **η^2^** |
| Intercept | 4.1 | 3.2 | 1.3 | .200 | - |
| SWI | -0.1 | 1.3 | -0.1 | .939 | < 0.01 |
| T1 ABC score | -3.5 | 2.0 | -1.7 | .092 | 0.07 |
| Age | 5.5 | 3.8 | 1.4 | .124 | 0.06 |
| Follow-up interval | -5.2 | 3.2 | -1.6 | .319 | 0.03 |
| Age x Follow-up interval | -3.8 | 3.0 | -1.3 | .212 | 0.04 |
| Longitudinal change in ABC scores ~ SWI + Timepoint 1 ABC score + Age + Follow-up interval + Age x Follow-up interval. R^2^ = .23, R^2^_adj_ = .12, F(5,34) = 2.1, *p* = .092. | | | | | |

| **Table S10:** General linear model showing relationship between small-world index, longitudinal changes in ABC scores, age and full-scale IQ in autistic 15-24-year-olds | | | | | |
| --- | --- | --- | --- | --- | --- |
| **N = 38 -** 15-24-year-olds | **Est. coeff.** | **Std. Error** | **t** | ***p*** | **η^2^** |
| Intercept | 8.0 | 3.0 | 2.7 | .011 |  |
| **SWI** | **-7.5** | **1.6** | **-4.8** | **< .001** | **0.18** |
| FSIQ | 4.1 | 1.3 | 3.1 | .004 | 0.07 |
| T1 ABC score | -8.0 | 1.4 | -5.8 | < .001 | 0.26 |
| Age | -2.1 | 2.8 | -0.7 | .466 | 0.04 |
| Follow-up interval | -1.3 | 2.7 | -0.5 | .629 | 0.11 |
| Sex (male) | -8.1 | 2.9 | -2.8 | .009 | 0.06 |
| Age x Follow-up interval | -8.5 | 3.5 | -2.4 | .021 | 0.05 |
| Longitudinal change in ABC score ~ **SWI** + FSIQ + Timepoint 1 ABC score + Age + Follow-up interval + Sex + Age x Follow-up interval. R^2^ = .67, R^2^_adj_ = .59, F(7,30) = 8.6, *p* < .001. | | | | | |
| Model statistics where SWI is omitted as a variable: R^2^ = .41, R^2^_adj_ = .29, F(6,31) = 3.6, *p* = .008. Change in R^2^_adj_ on adding SWI to model = +.30. | | | | | |

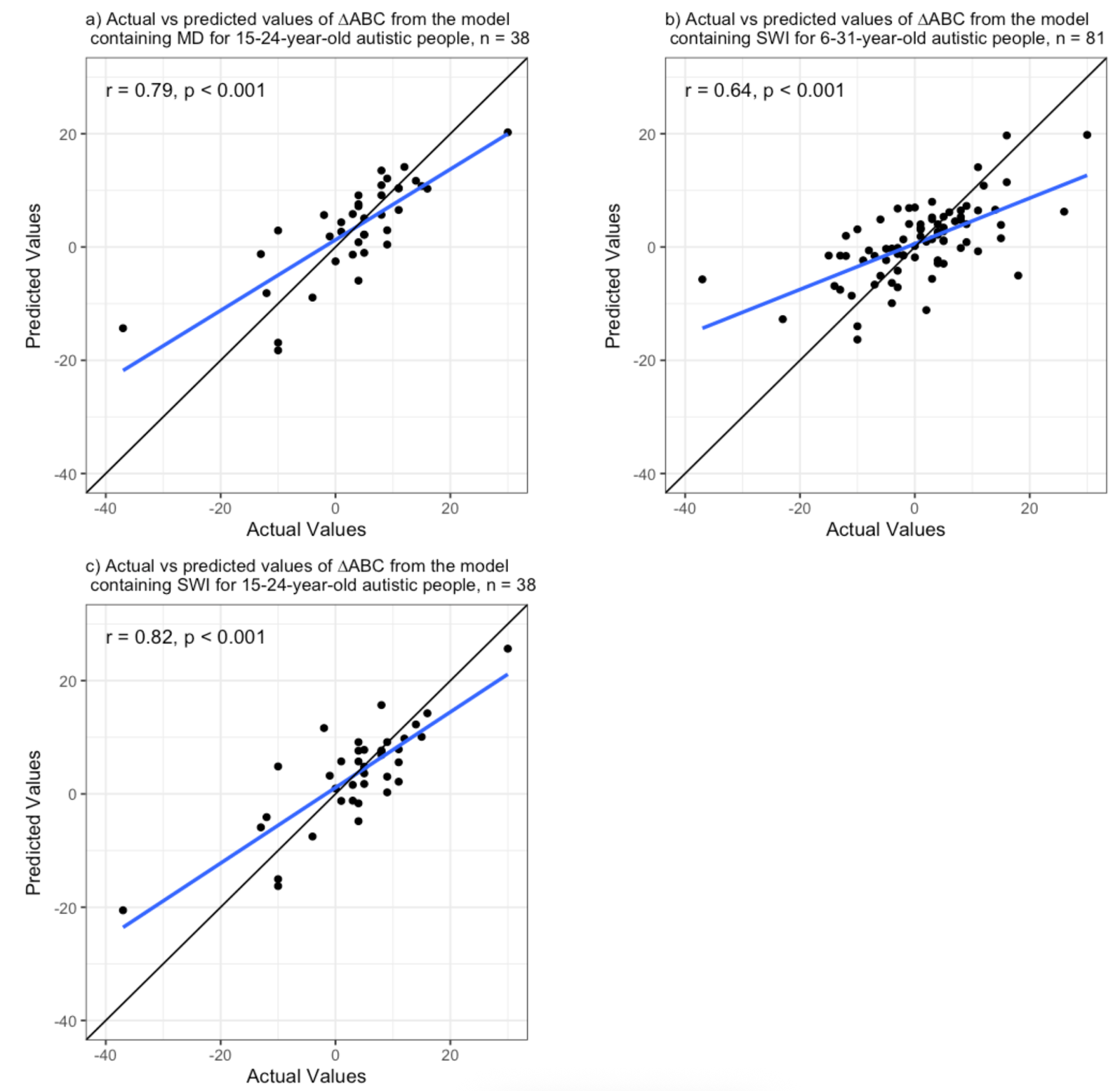

**Figure S6 – Associations between actual and predicted values of longitudinal changes in ABC scores.** Plots are interpreted as follows: gradient ~ 1: predicted values closely match observed values, gradient < 1: model underestimates true values, gradient > 1: model overestimates true values. The black lines are a slope of 1, while the blue lines are the regression line of the scatterplots. **a)** The model containing mean degree predicted longitudinal changes in ABC score in autistic 15-24-year-olds. Visual inspection shows the gradient of the regression line is approximately 1, suggesting that the model did not tend to either over- or under-estimate true values. **b)** The model containing small-world index significant predicted longitudinal changes in ABC score in autistic 6-31-year-olds. Visual inspection shows the gradient of the regression line is < 1, suggesting that the model tended to underestimate the magnitude of adaptive function changes. **c)** The model containing small-world index significantly predicted longitudinal changes in adaptive function in 15-24-year-old autistic people. Visual inspection shows the gradient of the regression line is approximately 1, suggesting that the model did not tend to either over- or under-estimate true values.

**Cross validation analysis for functional connectivity metrics significantly predicting longitudinal changes in ABC scores**

| **Table S11:** Cross-validation analysis for functional connectivity metrics significantly predicting longitudinal changes in ABC scores. 10-fold cross validation results of general linear models that predict adaptive function outcomes with a significant main effect of functional connectivity. High cross-validation R^2^s show that the models predict a substantial portion of variance in longitudinal changes in adaptive function, even in hold-out datasets. RMSE = root-mean-squared error; MAE = mean absolute error (range = -37 to + 30), SWI = small-world index, MD = mean degree, FSIQ = full-scale IQ. | | | | | |
| --- | --- | --- | --- | --- | --- |
|  | | | **10-fold cross validation:** | | |
| **Model** | **Age group** | **n** | **RMSE** | **R^2^** | **MAE** |
| ΔABC ~ **SWI** + FSIQ + T1 ABC score + Age + Follow-up interval + Site + Age x Follow-up interval | 6 - 31 yrs | 81 | 8.5 | .39 | 6.3 |
| ΔABC ~ **SWI** + FSIQ + T1 ABC score + Age + Follow-up interval + Sex + Age x Follow-up interval | 15 - 24 yrs | 38 | 7.7 | .62 | 6.1 |
| ΔABC ~ **MD** + FSIQ + T1 ABC score + Age + Follow-up interval + Sex + Site + Age x Follow-up interval | 15 - 24 yrs | 38 | 9.2 | .51 | 7.8 |

**The relationship between parent reported SRS T-score and functional connectivity metrics as a function of moving age**

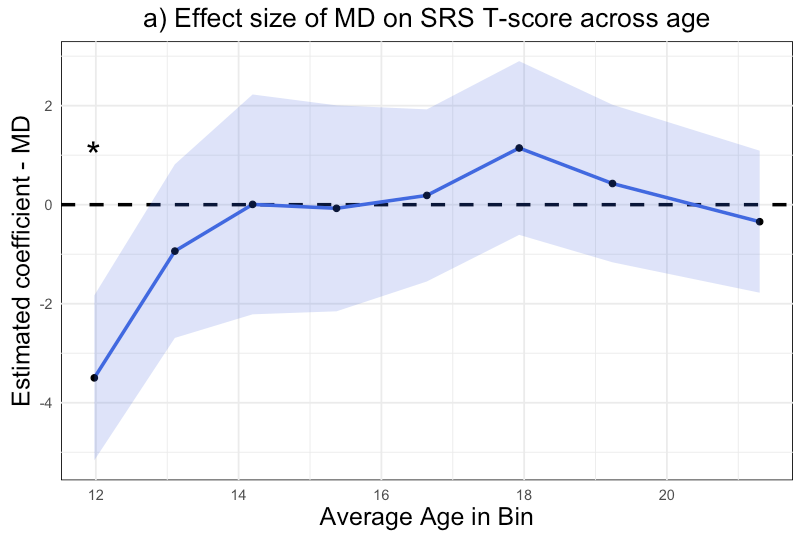

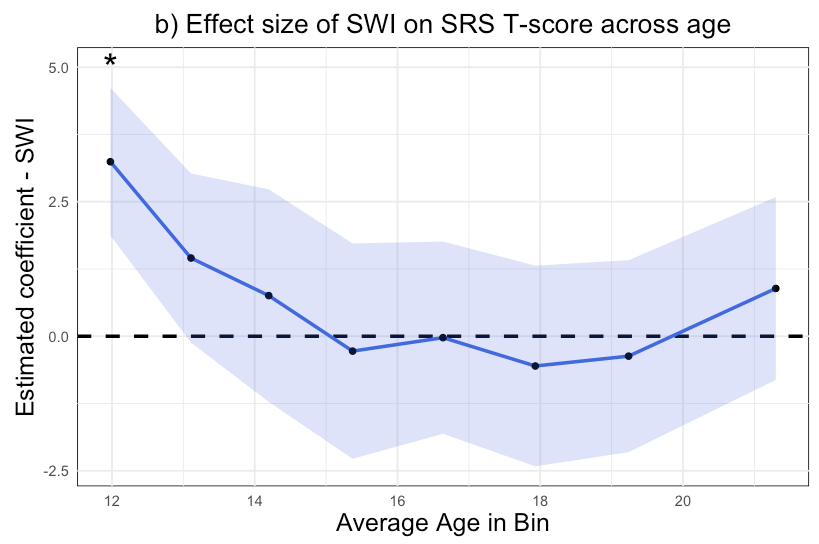

**Figure S7 –** Relationship between parent-reported SRS T-score and **a)** mean degree (MD) and **b)** small-world index (SWI) as a function of rolling average age. There was a main effect of MD/SWI in the youngest age-bracket (mean age = 12.0, minimum age = 7.9, maximum = 15.9 age years) only. The estimated coefficients for functional connectivity metrics were from general linear models containing site and age as covariates. To obtain estimated coefficients, we performed identical models in participant groups of n = 60, ordered by participant age, increasing by 8 participants at a time. * *p* < .05.

| **Table S12 – Moving estimated coefficients for mean degree and small-world index in GLMs predicting ΔABC, as a function of age.** Age brackets were *not* pre-defined, instead, successive GLMs (n = 30) were performed on participants of increasing age in steps of 5 participants. The GLM used was identical in form across ages and functional connectivity metric: ΔABC ~ Functional connectivity metric + FSIQ + Timepoint 1 ABC score + Age + Follow-up interval + Sex + Age x Follow-up interval. GLM: general linear model. Bold font: *p*_FDR_ < .05. | | | | | | | | |
| --- | --- | --- | --- | --- | --- | --- | --- | --- |
| **Age (years)** | | | **Mean Degree** | | | **Small-world index** | | |
| **Mean** | **Min.** | **Max.** | **Est. coefficient** | **Std. error** | ***p*_FDR_** | **Est. coefficient** | **Std. error** | ***p*_FDR_** |
| 10.5 | 7.9 | 13.3 | -0.6 | 1.9 | .751 | -1.5 | 1.7 | .597 |
| 11.5 | 8.7 | 14.3 | -1.5 | 1.9 | .751 | 0.0 | 1.9 | .998 |
| 12.5 | 9.9 | 15.0 | -1.1 | 2.8 | .751 | 0.0 | 1.9 | .998 |
| 13.4 | 10.8 | 16.6 | -0.6 | 1.8 | .751 | 1.0 | 1.4 | .683 |
| 14.4 | 11.3 | 17.4 | -1.0 | 1.7 | .751 | 1.7 | 1.3 | .339 |
| 15.4 | 12.8 | 17.9 | -1.3 | 1.8 | .751 | 2.0 | 1.3 | .288 |
| 16.3 | 13.4 | 19.4 | -1.2 | 1.6 | .751 | 1.9 | 1.2 | .288 |
| 17.4 | 14.5 | 20.9 | 0.6 | 1.5 | .751 | 0.3 | 1.4 | .986 |
| 18.6 | 15.2 | 23.0 | 5.0 | 1.9 | **.048** | -7.1 | 1.7 | **.001** |
| 19.9 | 16.7 | 24.2 | 7.1 | 2.0 | **.011** | -9.4 | 1.8 | **.000** |
| 21.8 | 17.5 | 30.3 | 6.2 | 1.8 | **.011** | -7.8 | 1.7 | **.001** |

**Relationship between functional connectivity metrics and longitudinal changes in adaptive function as a function of age**

**Comparison against other prognostic variables**

| **Table S13: comparing the utility of MD/SWI and behavioural measures in predicting longitudinal changes in adaptive function.** Initial models contain functional connectivity variables (small-world index or mean degree), and not the social-responsiveness scale (SRS) or full-scale IQ (FSIQ). When the functional connectivity variable is replaced with SRS or FSIQ in each model, the model R^2^_adj_ decreases, and there is no significant main effect of replacement variable. T1 = timepoint 1, ABC = adaptive behaviour composite score, SWI = small-world index, MD = mean degree, SRS = social-responsiveness scale, FSIQ = full-scale IQ. | | | | | | | |
| --- | --- | --- | --- | --- | --- | --- | --- |
| **Initial models** | | | **Replacement models, showing main effect of replacement variable** | | | | |
| **Dependent variable: ∆ABC**  **Independent variables:** | **Age group** | **Original variable of interest** | **Replaced with** | **Est. coeff.** | **Std. Error** | **∆Adj. R^2^** | ***p*** |
| **SWI** + T1 ABC + Age + Follow-up interval + Sex + Age x Follow-up interval | 15-24 | SWI | FSIQ | 3.89 | 1.96 | -.18 | .056 |
| **SWI** + T1 ABC + Age + Follow-up interval + Sex + Age x Follow-up interval | 15-24 | SWI | SRS | -2.89 | 3.12 | -.31 | .364 |
| **SWI** + T1 ABC + Age + Follow-up interval + Site + Age x Follow-up interval | 6-31 | SWI | FSIQ | 1.28 | 1.14 | -.04 | .261 |
| **SWI** + T1 ABC + Age + Follow-up interval + Site + Age x Follow-up interval | 6-31 | SWI | SRS | -2.68 | 1.65 | -.06 | .105 |
| **MD** + T1 ABC + Age + Follow-up interval + Sex + Site + Age x Follow-up interval | 15-24 | MD | FSIQ | 4.31 | 2.00 | -.07 | .040 |
| **MD** + T1 ABC + Age + Follow-up interval + Sex + Site + Age x Follow-up interval | 15-24 | MD | SRS | -1.96 | 3.61 | -.26 | .592 |

To investigate the relative utility of MD and SWI as prognostic biomarkers of adaptive function outcomes, we compared their predictive performance to that of T1 autistic features (SRS T-score) and T1 full-scale IQ (FSIQ). For the models with a significant main effect of functional connectivity metric on adaptive function outcomes, we compared models that contained a functional connectivity variable (without SRS T-score or FSIQ) with models where the functional connectivity variable was *replaced* by SRS T-score or FSIQ. Models containing SRS or FSIQ instead of the functional connectivity variable led to decreased R^2^_adj_, with no main effect of SRS or FSIQ (Table S13). Then, we compared models that contained SRS or FSIQ *in addition* to the functional connectivity metric. Adding FSIQ increased the R^2^_adj_ and decreased the AIC of the respective models (despite no main effect of FSIQ); thus, FSIQ was included in the main models. Adding SRS decreased the R^2^_adj_ and increased the AIC of the models.

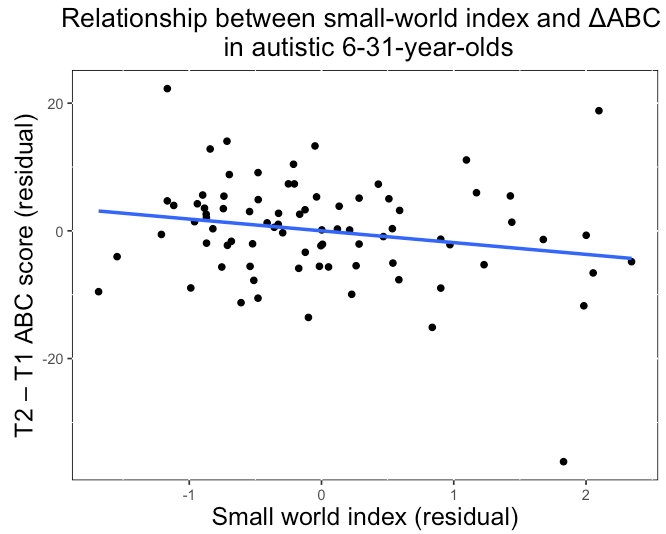

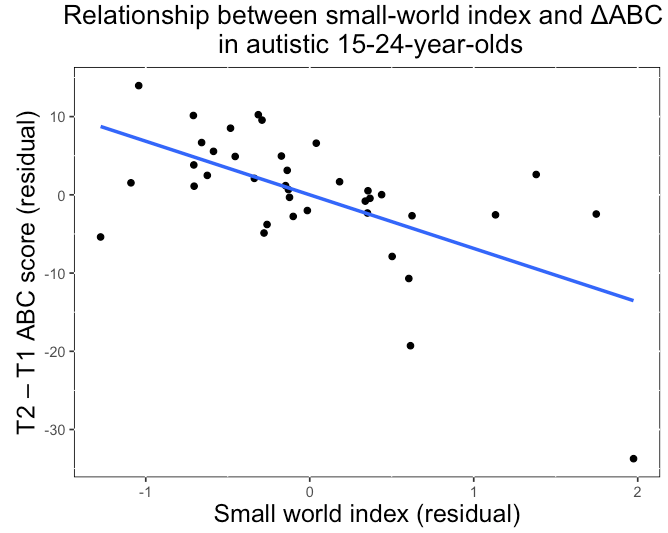

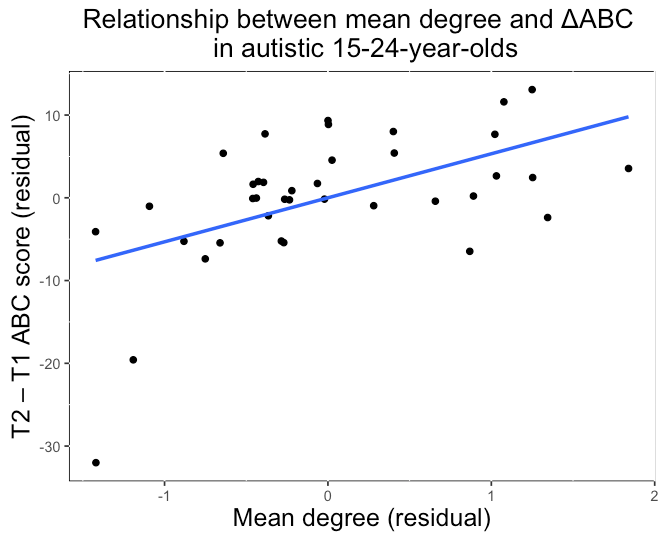

partial *r* = .53, *p* = .001

partial *r* = -.60, *p* < .001

e)

partial *r* = -.21, *p* = .062

c)

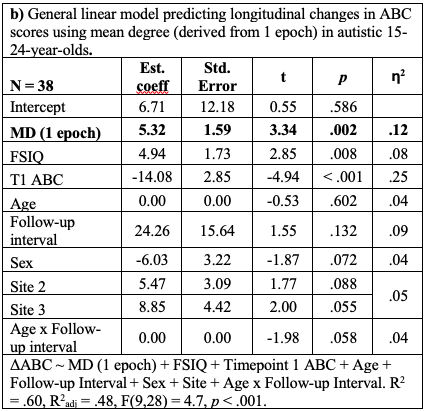

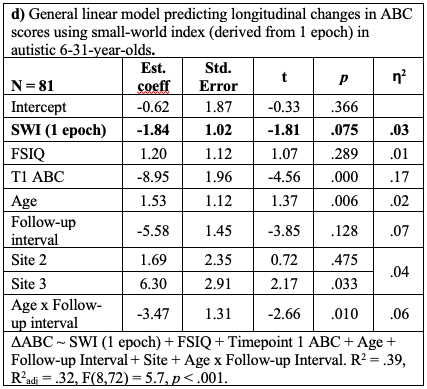

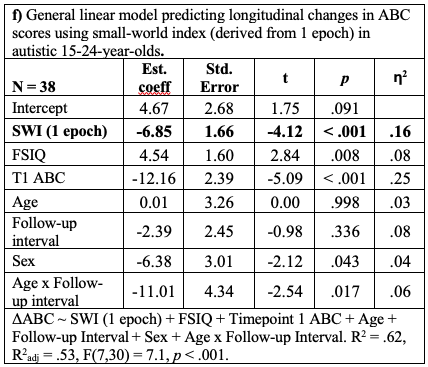

**Figure S8 – Relationship between longitudinal changes in adaptive function in autistic people and functional connectivity metrics derived from only one 20-second EEG epoch**. The associations between the functional connectivity metrics and adaptive function changes were similar to values derived from the mean of one-to-four epochs (as in the main analysis), although expectedly smaller in magnitude. The values in the graphs **a)**, **c)** and **e)** are residualised for the co-variates listed in **b)**, **d)** and **f)** respectively**.** ΔABC: change in adaptive behaviour composite score.

**Polygenic scores**

| **Table S14**- General linear model showing relationship between mean degree, polygenic scores for autism, age and sex | | | | | |
| --- | --- | --- | --- | --- | --- |
| **N = 136** | **Est. coeff** | **Std. Error** | **t** | **p** | **η^2^** |
| Intercept | 22.35 | 4.08 | 5.47 | < .001 | - |
| **PGS autism** | 0.03 | 0.45 | 0.06 | .95 | .00 |
| Age | 1.23 | 0.49 | 2.50 | .01 | .04 |
| Age^2^ | -0.02 | 0.01 | -1.82 | .07 | .02 |
| Sex (male) | 1.36 | 1.06 | 1.29 | .20 | .01 |
| MD ~ polygenic score for autism + age + age^2^ + sex. R^2^ = .16, R^2^_adj_ = .14, F(4,131) = 6.44, *p* < .001. | | | | | |

| **Table S15** - General linear model showing relationship between small-world index, polygenic scores for autism, age and sex | | | | | |
| --- | --- | --- | --- | --- | --- |
| **N = 136** | **Est. coeff** | **Std. Error** | **t** | **p** | **η^2^** |
| Intercept | 1.14 | 0.04 | 26.24 | < .001 | - |
| **PGS autism** | 0.00 | 0.00 | -0.42 | .67 | .00 |
| Age | -0.01 | 0.01 | -2.67 | .01 | .05 |
| Age^2^ | 0.00 | 0.00 | 2.16 | .03 | .03 |
| Sex (male) | -0.01 | 0.01 | -0.87 | .39 | .01 |
| SWI ~ polygenic score for autism + age + age^2^ + sex. R^2^ = .12, R^2^_adj_ = .09, F(4,131) = 4.5, *p* = .002. | | | | | |

| **Table S16** – Variances of PGS and functional connectivity metrics were similar in autistic and non-autistic groups | | |
| --- | --- | --- |
| **Measure** | **Variance in autistic people** | **Variance in non-autistic people** |
| PGS total brain volume | 0.92 | 1.12 |
| Mean degree | 34.5 | 34.5 |
| Small-world index | 0.004 | 0.004 |

**No associations between longitudinal changes in ABC scores and head circumference**

We examined models predicting longitudinal changes in adaptive function with the variable head circumference either added to or used instead of the functional connectivity variable. There was no main effect of head circumference on ABC score changes in either case. Further, associations between PGS and connectivity metrics remained significant even when adjusting for head circumference (Tables S17-19).

| **Table S17** – general linear model showing that associations between SWI and longitudinal changes in ABC scores are still significant when head circumference is added as a predictor variable, in autistic people aged 6-31-years. | | | | | |
| --- | --- | --- | --- | --- | --- |
|  | **Est. coeff** | **Std. Error** | **t value** | **p** | **η^2^** |
| Intercept | 7.84 | 24.10 | 0.33 | .746 |  |
| **SWI (standardised)** | **-2.62** | **1.07** | **-2.45** | **.017** | **0.05** |
| FSIQ | 0.09 | 0.07 | 1.42 | .161 | 0.02 |
| Head Circumference | -0.28 | 0.25 | -1.09 | .282 | 0.01 |
| T1 ABC score | -0.48 | 0.10 | -4.67 | < .001 | 0.18 |
| Age | 3.67 | 1.24 | 2.95 | .004 | 0.01 |
| Age difference | 0.05 | 0.03 | 1.71 | .092 | 0.06 |
| Site 2 | 1.21 | 2.49 | 0.49 | .628 | 0.04 |
| Site 3 | 6.32 | 3.13 | 2.02 | .047 |  |
| Age x age difference | -0.01 | 0.00 | -2.78 | .007 | 0.07 |
| Longitudinal changes in ABC score ~ SWI (standardised) + FSIQ + T1 ABC score + Head circumference Age + Age difference + Site + age x age difference. R^2^ = .43, R^2^_adj_ = .35, F(9,67) = 5.6, *p* < .001. | | | | | |

| **Table S18** – general linear model showing that associations between SWI and longitudinal changes in ABC scores are still significant when head circumference is added as a predictor variable, in autistic people aged 15-21-years. | | | | | |
| --- | --- | --- | --- | --- | --- |
|  | **Est. coeff** | **Std. Error** | **t value** | **p** | **η^2^** |
| Intercept | -45.23 | 194.80 | -0.23 | .819 |  |
| **SWI (standardised)** | **-5.92** | **1.58** | **-3.74** | **.002** | **0.22** |
| FSIQ | 0.20 | 0.12 | 1.67 | .113 | 0.04 |
| Head Circumference | -0.45 | 0.25 | -1.84 | .083 | 0.05 |
| T1 ABC score | -0.57 | 0.19 | -2.93 | .009 | 0.14 |
| Age | 7.07 | 10.90 | 0.65 | .525 | 0.14 |
| Age difference | 0.22 | 0.32 | 0.68 | .506 | 0.10 |
| Sex (male) | -5.12 | 4.04 | -1.27 | .223 | 0.03 |
| Age x age difference | -0.01 | 0.02 | -0.85 | .407 | 0.01 |
| Longitudinal changes in ABC score ~ SWI (standardised) + FSIQ + T1 ABC score + Head circumference Age + Age difference + Site + age x age difference. R^2^ = 71, R^2^_adj_ = .58, F(8,17) = 5.3, *p* = .002. | | | | | |

| **Table S19** – general linear model showing that associations between SWI and longitudinal changes in ABC scores are still significant when head circumference is added as a predictor variable, in autistic people aged 15-21-years. | | | | | |
| --- | --- | --- | --- | --- | --- |
|  | **Est. coeff** | **Std. Error** | **t value** | **p** | **η^2^** |
| Intercept | -171.50 | 182.20 | -0.94 | .361 |  |
| **MD (standardised)** | **5.51** | **1.43** | **3.86** | **.002** | **0.18** |
| FSIQ | 0.17 | 0.11 | 1.46 | .164 | 0.03 |
| Head Circumference | -0.33 | 0.23 | -1.43 | .172 | 0.02 |
| T1 ABC score | -0.66 | 0.19 | -3.53 | .003 | 0.15 |
| Age | 0.04 | 0.03 | 1.36 | .194 | 0.22 |
| Age difference | 0.43 | 0.30 | 1.45 | .168 | 0.10 |
| Sex (male) | -3.27 | 3.67 | -0.89 | .387 | 0.01 |
| Site 2 | 5.31 | 3.10 | 1.72 | .107 | 0.09 |
| Site 3 | 11.37 | 4.52 | 2.52 | .024 |  |
| Age x age difference | 0.00 | 0.00 | -1.66 | .119 | 0.03 |
| Longitudinal changes in ABC score ~ MD (standardised) + FSIQ + T1 ABC score + Head circumference Age + Age difference + Site + age x age difference. R^2^ = 78, R^2^_adj_ = .64, F(10,15) = 5.5, *p* = .002. | | | | | |

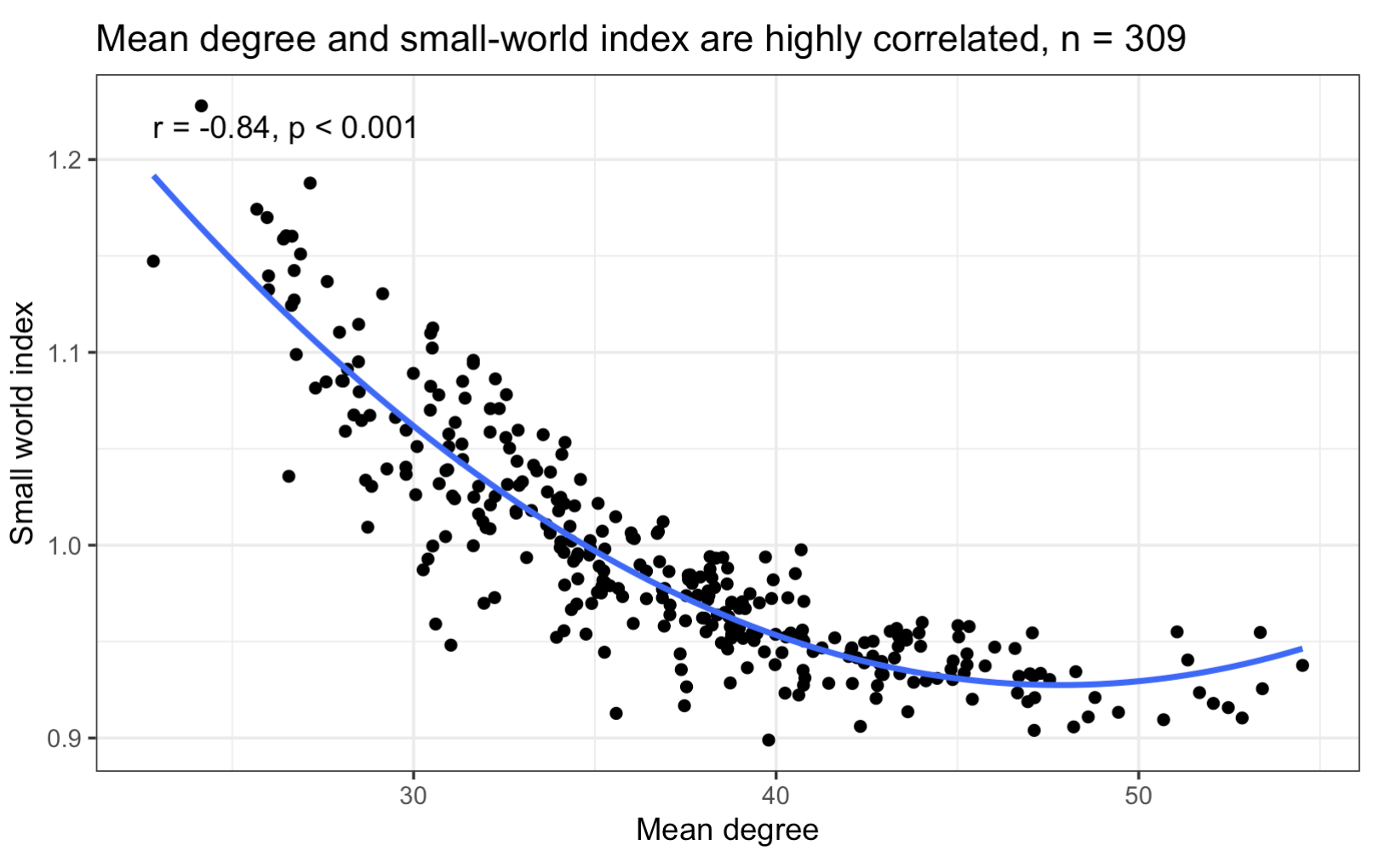

**Figure S9 –** Mean degree strongly negatively correlates with small-world index.

**Sensitivity analysis of functional connectivity predicting adaptive function outcomes in participants without ADHD**

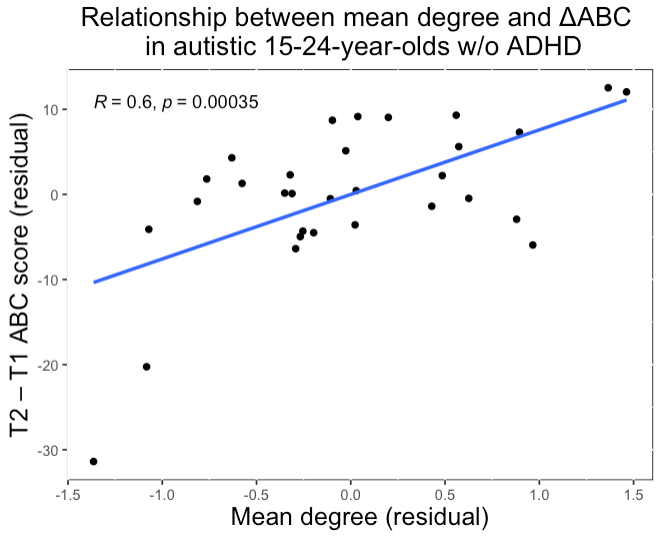

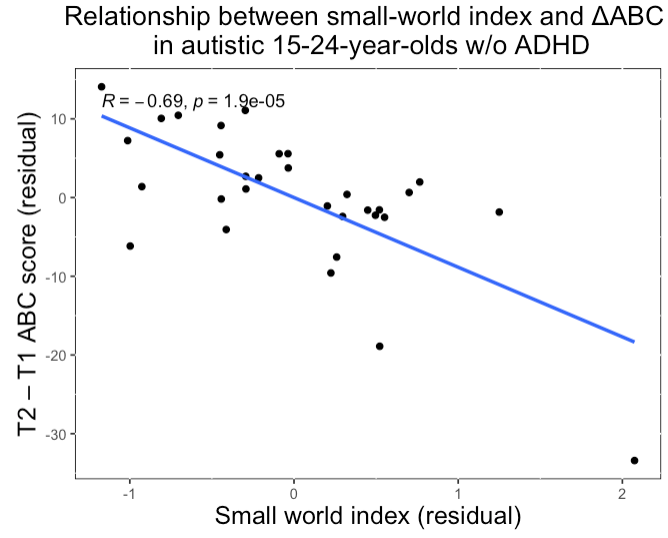

**Figure S10** – Relationships between functional connectivity metrics and longitudinal changes in ABC scores remained consistent in a sample of autistic people without ADHD, n = 31.
